# Epidemiological cutoff values for a 96-well broth microdilution plate for high-throughput research antibiotic susceptibility testing of *M. tuberculosis*

**DOI:** 10.1101/2021.02.24.21252386

**Authors:** The CRyPTIC Consortium, Philip W Fowler, Ivan Barilar, Simone Battaglia, Emanuele Borroni, Angela Pires Brandao, Alice Brankin, Andrea Maurizio Cabibbe, Joshua Carter, Daniela Maria Cirillo, Pauline Claxton, David A Clifton, Ted Cohen, Jorge Coronel, Derrick W Crook, Viola Dreyer, Sarah G Earle, Vincent Escuyer, Lucilaine Ferrazoli, George Fu Gao, Jennifer Gardy, Saheer Gharbia, Kelen Teixeira Ghisi, Arash Ghodousi, Ana Luíza Gibertoni Cruz, Louis Grandjean, Clara Grazian, Ramona Groenheit, Jennifer L Guthrie, Wencong He, Harald Hoffmann, Sarah J Hoosdally, Martin Hunt, Zamin Iqbal, Nazir Ahmed Ismail, Lisa Jarrett, Lavania Joseph, Ruwen Jou, Priti Kambli, Rukhsar Khot, Jeff Knaggs, Anastasia Koch, Donna Kohlerschmidt, Samaneh Kouchaki, Alexander S Lachapelle, Ajit Lalvani, Simon Grandjean Lapierre, Ian F Laurenson, Brice Letcher, Wan-Hsuan Lin, Chunfa Liu, Dongxin Liu, Kerri M Malone, Ayan Mandal, Mikael Mansjö, Daniela Matias, Graeme Meintjes, Flávia de Freitas Mendes, Matthias Merker, Marina Mihalic, James Millard, Paolo Miotto, Nerges Mistry, David Moore, Kimberlee A Musser, Dumisani Ngcamu, Hoang Ngoc Nhung, Stefan Niemann, Kayzad Soli Nilgiriwala, Camus Nimmo, Nana Okozi, Rosangela Siqueira Oliveira, Shaheed Vally Omar, Nicholas Paton, Timothy EA Peto, Juliana Maira Watanabe Pinhata, Sara Plesnik, Zully M Puyen, Marie Sylvianne Rabodoarivelo, Niaina Rakotosamimanana, Paola MV Rancoita, Priti Rathod, Esther Robinson, Gillian Rodger, Camilla Rodrigues, Timothy C Rodwell, Aysha Roohi, David Santos-Lazaro, Sanchi Shah, Thomas Andreas Kohl, Grace Smith, Walter Solano, Andrea Spitaleri, Philip Supply, Utkarsha Surve, Sabira Tahseen, Nguyen Thuy Thuong Thuong, Guy Thwaites, Katharina Todt, Alberto Trovato, Christian Utpatel, Annelies Van Rie, Srinivasan Vijay, Timothy M Walker, A Sarah Walker, Robin Warren, Jim Werngren, Maria Wijkander, Robert J Wilkinson, Daniel J Wilson, Penelope Wintringer, Yu-Xin Xiao, Yang Yang, Zhao Yanlin, Shen-Yuan Yao, Baoli Zhu

## Abstract

Drug susceptibility testing of *M. tuberculosis* is rooted in a binary susceptible/resistant paradigm.

Whilst there are considerable advantages in measuring the minimum inhibitory concentrations (MICs) of a panel of drugs for an isolate it is necessary to measure the epidemiological cutoff values (ECOFF/ECVs) to permit comparison with qualitative data. Here we present ECOFF/ECVs for 13 anti-TB compounds, including bedaquiline and delamanid, derived from 20,637 clinical isolates collected by 14 laboratories based in 11 countries on five continents. Each isolate was incubated for 14 days on a dry 96-well broth microdilution plate and then read. Resistance to most of the drugs due to prior exposure is expected and the MIC distributions for many of the compounds are complex and therefore a phenotypically wild-type population could not be defined. Since a majority of samples also underwent genetic sequencing, we defined a genotypically wild-type population and measured the MIC of the 99^th^ percentile by direct measurement and via fitting a Gaussian using interval regression.

The proposed ECOFF/ECV values were then validated by comparing to the MIC distributions of high-confidence genetic variants that confer resistance and to qualitative drug susceptibility tests obtained via Mycobacterial Growth Indicator Tube and the Microscopic-Observation Drug-Susceptibility assay.

These ECOFF/ECV values will inform and encourage the more widespread adoption of broth microdilution – this is a cheap culture-based method that tests the susceptibility of 12-14 antibiotics on a single 96-well plate and so could help personalise the treatment of tuberculosis.

## INTRODUCTION

*Mycobacterium tuberculosis* kills more people worldwide than any other single pathogen, SARS-CoV-2 excepted (1). Despite its impact on global health, antibiotic susceptibility testing (AST) *for M. tuberculosis*, has lagged behind other bacterial diseases due to its slow growth rate, difficulty in culturing and its low prevalence in high-income countries. The consequence is that most patients in the world receive empiric, or semi-empiric, treatment, which reduces the chance of treatment success and risks the amplification of resistance where too few effective drugs are prescribed.

The dramatic reduction in genetic sequencing costs has enabled genetics-based AST where the genome of a pathogen is sequenced and then examined for known variants that confer resistance to specific antibiotics. *M. tuberculosis* is well-suited to this approach (2–10), and several public health bodies have adopted whole genome sequencing as their standard AST method (11). Although PCR platforms can deliver universal antibiotic susceptibility testing in its narrowly defined sense (12), genome sequencing is the only approach that can realistically deliver comprehensive AST in settings where phenotyping remains too expensive and too infrastructure dependent, and comprehensive AST is the only way to optimise treatment regimens and outcomes.

The Comprehensive Research Prediction for Tuberculosis: an International Consortium (CRyPTIC) research project has collected 20,637 clinical *M. tuberculosis* samples from across the world. The primary aim of the project is to identify mutations in the *M. tuberculosis* genome that confer phenotypic resistance to a wide range of antibiotics. The CRyPTIC project measured minimum inhibitory concentrations (MIC) of each drug to permit quantitative analyses, associating mutations with MIC values with a view to using genome sequencing data to personalise drug regimens and doses. From the start the CRyPTIC project has taken a data-driven approach whereby all analyses are algorithmic, hence the allocation of a sample to subgroup requires little or no expert, and hence subjective, intervention. This has the virtue of ensuring the results are reproducible.

The most practical and affordable means of determining MICs at scale was to use a pre-prepared 96-well 7H9 broth microdilution plate based on the Thermo Fischer Sensititre MYCOTB MIC plate (13–18), but including the new or repurposed antibiotics that feature in current WHO guidance (19). The CRyPTIC project designed a variant of the MYCOTB plate, called UKMYC5, that contains fourteen antibiotics, including bedaquiline, delamanid, clofazimine and linezolid but not pyrazinamide (Fig. 1A). Based on a multi-laboratory study that examined the inter- and intra-laboratory reproducibility of the UKMYC5 plate and determined the optimum reading methods and incubation period (17), CRyPTIC subsequently modified the design by removing para-aminosalicylic acid and extending/changing the concentration of certain drugs, leading to the 13-drug UKMYC6 plate (Fig. 1B).

**Figure 1.**
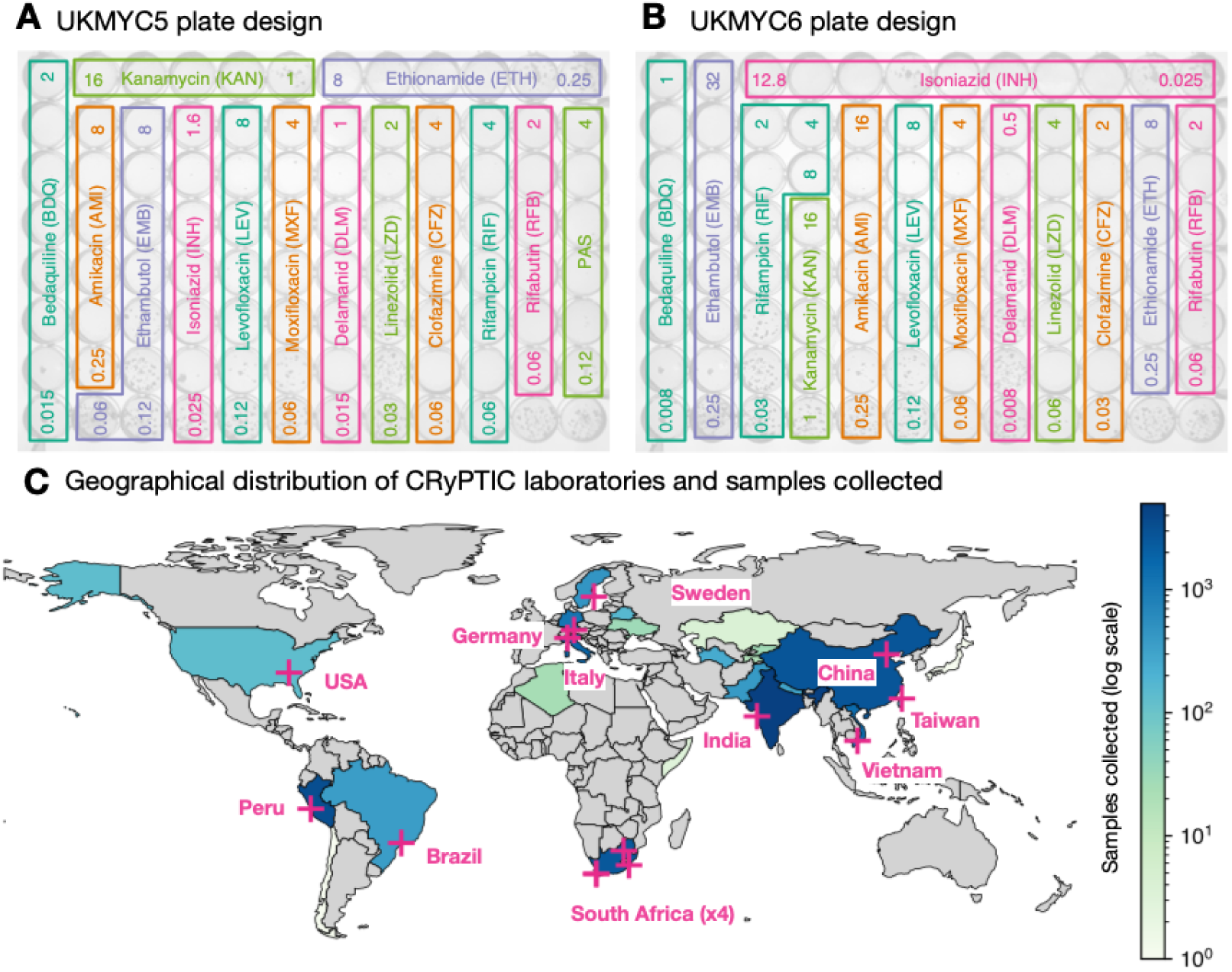
The CRyPTIC consortium has collected 20,637 clinical tuberculosis samples worldwide. The layout and concentrations of the anti-TB drugs on the (**A**) UKMYC6 and (**B**) UKMYC5 microdilution 96-well plates. All concentrations are in mg/L and for clarity only the first and last concentration in each doubling series are given. The two unlabelled wells in the bottom right-hand corner contain no antibiotic and are therefore positive controls. Note that all doubling As expected, the MIC histograms differ between drugs (Fig. 2, S5, S6); the MICs for some compounds form bimodal distributions (INH, KAN, AMI, RIF, RFB) and therefore conform to the classical binary paradigm whereby an isolate is either ‘resistant’ or ‘susceptible’. CRyPTIC aimed for half the isolates collected to be multi-drug resistant (MDR) and the MIC histograms for isoniazid and rifampicin are consistent with this. Given this bias towards MDR in the dataset, one would expect appreciable resistance to ethambutol, ethionamide, both fluoroquinolones and both aminoglycosides. Both drugs belonging to the latter class indeed have a subset of isolates with very high MICs. The MIC histograms for the remaining compounds (EMB, ETH, MXF, LEV) are not bimodal hence it is unclear whether they can be adequately described by two log-normal distributions. Since the remaining drugs on the plates (BDQ, DLM, CFZ, LZD) have not yet been widely used, and for some countries were not even available to treat tuberculosis, one expects little resistance in the dataset and hence it is likely these MIC histograms are ‘phenotypically wild-type’ (pWT). All the MIC histograms are truncated/censored at either one or both ends, and some are severely truncated with the mode MIC occurring in the lowest dilution (AMI, RFB, DLM). Our large dataset allows us to use reproducible, algorithmic approaches for estimating the 99^th^ percentile of the wild-type population.

In this paper we propose epidemiological cut-off values (ECVs or ECOFFs) for the UKMYC series of plates to enable subsequent research on this dataset. The ECOFF is the highest MIC observed within a phenotypically wild-type population, usually defined as the MIC which encompasses 99% of that population (20), and allows interpretation of an MIC value as ‘susceptible or ‘resistant’ – crucial to the decision on whether to prescribe a drug. The standard approach requires uncensored MICs and assumes that the phenotypic wild-type population can be readily identified, either because the population has been minimally exposed to the drug, or because the MIC distribution is strongly bimodal. These conditions are not universally met in our dataset and we shall therefore identify a *genotypically* wild-type population from which we can either measure the ECOFF/ECV directly or via a Gaussian fitted using interval regression, a statistical technique that can fit to censored data. We have made Python code publicly available that enables anyone to reproduce most of the figures and tables in a web browser window (21). Although ECOFF/ECVs have been proposed for the MYCOTB microdilution plate using 385 strains from South Africa (22), we are here able to draw upon a far larger and more geographically diverse *M. tuberculosis* dataset.

## RESULTS

AST was performed on 20,637 isolates to 13 anti-TB drugs using either the UKMYC6 (12,672, 61%) or the UKMYC5 (7,965, 39%) plate design (Table 1, Fig. 1A & B). These data were generated in fourteen CRyPTIC laboratories based in eleven countries on five continents (Fig. 1C, Table S1). The isolates themselves were collected from 27 countries, with 19 countries contributing ten or more, and 15 countries contributing 100 or more isolates (Table S2). Due to differences between the laboratories, it was not possible to collect clinical outcome data for the samples. Quality control processes detected that one laboratory developed a problem inoculating the plates – these plates were removed – and that another laboratory never managed to inoculate successfully: all their plates were excluded. Excluded these left 17,054 plates.

**Table 1.** The number of isolates collected, split by the two microtitre plate designs used. An asterisk indicates that this is the average number of plates across all drugs. This table can be reproduced (21).

|  | Total | UKMYC6 | UKMYC5 |
| --- | --- | --- | --- |
| Isolates collected | 20,637 | 12,672 | 7,965 |
| Readable plates | 17,054 | 10,010 | 7,044 |
| Readable plates with images | 15,138 | 9,272 | 5,866 |
| Readable plates with genetics | 12,362 | 6,019 | 6,343 |
| Readable plates with genetics and images | 10,938 | 5,552 | 5,386 |
| Readable plates with images and passing quality assurance | *11,801 | *6,896 | *4,904 |
| Readable plates with genetics, images and passing quality assurance | *8,553 | *4,027 | *4,526 |
| Readable plates with genetic, images, passing quality assurance and genotypically wild-type | *3,328 | *1,606 | *1,722 |

Of these 12,362 also had their whole genome sequenced (Methods, Table 1), allowing us to infer species and lineage information using SNP-IT (23). All isolates belonged to the *Mycobacterium tuberculosis* complex (MBTC), with the majority (12,348, 99.9%) confirmed as *M. tuberculosis* (Table S3), of which the majority belonged to either Lineage 2 (35%) or 4 (50%, Table S4) with the expected geographic distribution (Table S5, Fig. S1) (24).

A previous study demonstrated that MICs measured by a single laboratory scientist after 14 days incubation of the UKMYC5 plate using either a Thermo Fisher Vizion instrument or a mirrored-box were reproducible and accurate (17). As a further reproducibility check we pooled the MIC measurements of the H37Rv reference strain that were taken as part of our quality control process (Fig. S2, Table S6); the histograms for both UKMYC plates showed that the majority of MICs measured by the laboratory scientists for many, but not all, of the drugs lay within one doubling dilution of the mode. Since the magnitude of MIC measurement error is anticipated to be much greater than the error in the genetic sequencing, we constructed an MIC quality assurance (QA) process to minimize the measurement error of the MICs (Fig. S3). This measured each MIC using up to three independent methods and only MICs where two of these methods concur are allowed into the final dataset. Overall, 77% of all MIC measurements passed the MIC QA process.

### The MIC histograms are different for different drugs

#### Iteratively fitting a log-normal distribution

ECOFFinder is a heuristic approach that attempts to iteratively fit a log-normal distribution to the MIC histogram and is recommended by both EUCAST (20, 25) and the CLSI. EUCAST advise that ECOFFinder should not be applied to truncated data, but we here we apply it to demonstrate how it performs for different levels of censored data. Distributions derived using ECOFFinder (Fig. 2, S6 & Table 2) describe our data well where the MIC histogram is minimally truncated (ETH, LZD, BDQ), however where the MIC histogram is heavily truncated (AMI, RFB, DLM) the resulting log-normal distribution does not fit the MIC histogram, and where the mode MIC is resistant (RIF) it fails to perform a fit at all. In addition, since ECOFFinder requires a single consistent MIC distribution and our dataset is composed of two plate designs, two ECOFF/ECVs are returned for each drug. For many drugs these are very similar but for ethambutol and delamanid the estimates are almost a doubling dilution different.

**Table 2.** The 99^th^ percentiles of the wild-type population as determined by three different algorithmic approaches and the resulting proposed ECOFF/ECVs for the thirteen drugs on the UKMYC6/5 plates.

| Drug |  | ECOFFfinder<br>(mg/L) |  | Direct measurement on<br>the gWT (mg/L) |  | Interval regression<br>on gWT (mg/L) | Proposed<br>ECOFF/<br>ECVs<br>(mg/L) |
| --- | --- | --- | --- | --- | --- | --- | --- |
|  |  | UKMYC6 | UKMYC5 | UKMYC6 | UKMYC5 | Both |  |
| Isoniazid | INH | 0.06 | - | 1.6 | 0.1 | 0.25 | 0.1 |
| Rifampicin | RIF | - | 0.08 | 0.5 | 0.25 | 0.52 | 0.5 |
| Ethambutol | EMB | 2.1 | 4.0 | 4 | 4 | 3.7 | 4 |
| Moxifloxacin | MXF | 0.74 | 0.75 | 1 | 2 | 1.4 | 1 |
| Levofloxacin | LEV | 0.63 | 0.72 | 1 | 4 | 1.3 | 1 |
| Kanamycin | KAN | 3.1 | 2.9 | 8 | 4 | 7.0 | 4 |
| Amikacin | AMI | 0.34 | 0.31 | 1 | 1 | 1.6 | 1 |
| Ethionamide | ETH | 3.1 | 3.0 | 4 | 4 | 4.0 | 4 |
| Rifabutin | RFB |  | - | 0.12 | 0.12 | 0.09 | 0.12 |
| Clofazimine | CFZ | 0.12 | 0.078 | 0.25 | 0.5 | 0.34 | 0.25 |
| Linezolid | LZD | 0.81 | 0.95 | 1 | 1 | 1.6 | 1 |
| Delamanid | DLM | 0.010 | 0.019 | 0.12 | 0.12 | 0.11 | 0.12 |
| Bedaquiline | BDQ | 0.11 | 0.11 | 0.25 | 0.25 | 0.20 | 0.25 |

**Figure 2.**
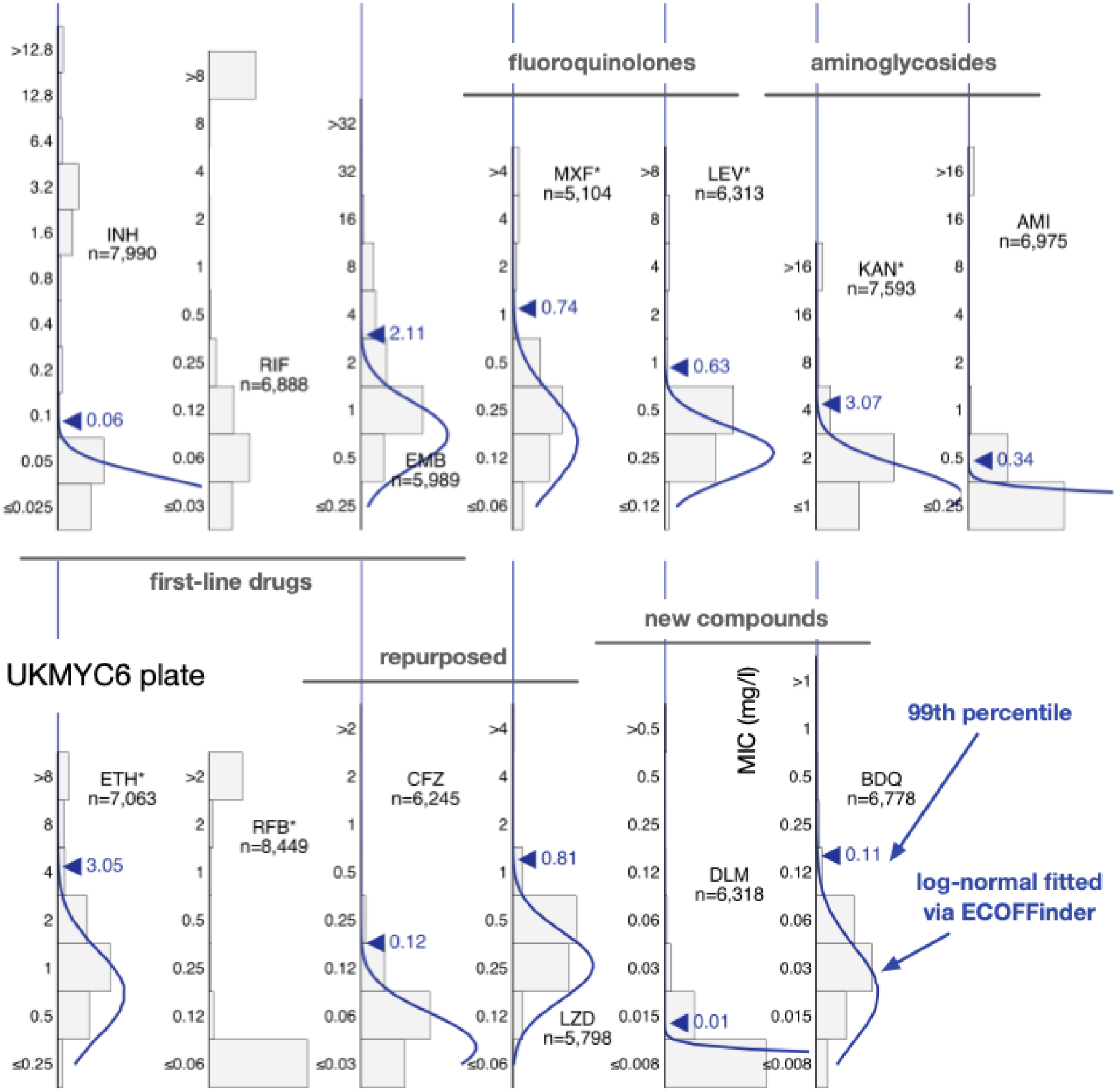
The MIC histograms for the 13 antibiotics on the UKMYC6 plate. Only MICs which have passed the quality assurance process described in the Methods are shown. ECOFFinder was used to fit a log-normal distribution to each histogram; this is drawn in blue and the resulting 99^th^ percentile is labelled. ECOFFinder was unable to fit a log-normal to both rifampicin (RIF) and rifabutin (RFB). See Fig. S6 for the UKYMYC5 histograms and the Supplemental Information for the numerical data. The histograms can be reproduced online (21).

To overcome the problem that our MIC histograms are truncated we applied *interval regression –* an established statistical method for fitting normal distributions to truncated data (26, 27) which, unlike conventional maximum-likelihood algorithms, takes into account that observations are properly represented by *intervals*. The entire dataset containing measurements from both plate designs can then be considered simultaneously, resulting in a single pair of log-normal distributions that describe the MIC histograms on both plate designs (Fig. S7). The model fails to converge for kanamycin and ethambutol and for several drugs the second distribution has a variance much larger than the MIC range which is nonsensical (AMI, ETH, RFB, CFZ, LZD, DLM, BDQ), although for the new- and repurposed compounds this is understandable since we do not expect many resistant isolates. Where the two distributions describe the data reasonably well (INH, RIF, MXF, LEV), they are well-separated, as defined by the 99^th^ percentile of the lower distribution (ECOFF/ECV) being smaller than the 1^st^ percentile of the upper distribution (the non-wild-type cut-off value, NCOFF), with the exception of isoniazid where NCOFF < ECOFF.

#### Defining a *genotypically wild-type* population

Using these approaches we were not able to produce acceptable results when the MIC histogram is truncated and/or is not clearly bimodal. In the latter case it is probable that the overall MIC histogram is a convolution of several smaller, narrower distributions. Genetics offers a way to disentangle these sub-populations: one can predict genetically the susceptibility of strains to most, but not all, of the 13 anti-TB compounds of interest (6–8). Note that we were unable to use the newer and more comprehensive genetic catalogue released by the WHO since its derivation set included these samples (9, 10). We predicted the antibiogram for the first-line (INH, RIF, EMB and also PZA – see below) and second-line (AMI, KAN, LEV, MXF, ETH) compounds (Methods). No predictions were made for the other anti-TB compounds on the plate since the association between genetics and their resistance is poorly understood at present.

We defined an isolate as being *genotypically wild-type* (gWT) if it is predicted to be susceptible to the four first-line compounds and not resistant to the second-line compounds (see ref (6) for the distinction). The laxer criterion for the second-line compounds allowed for the fact that our understanding for these drugs is less complete. Epidemiologically, it is the case that if an isolate is susceptible to all four first-line antibiotics, it is also likely to be susceptible to second-line antibiotics (except perhaps for prior fluoroquinolone exposure or deeply rooted second-line resistance mutations). To contribute a laboratory had to have collected susceptible samples which had undergone whole genome sequencing and also had a high quality photograph taken of the UKMYC plate after 14 days incubation so we could run the QA process. As a result isolates from only nine CRyPTIC laboratories made up this dataset (Table S8) and the number of confirmed MICs varied between 2,594 and 4,078 by drug, with a mean of 3,263 (Table S9).

Visually, the resulting MIC histograms are simpler and more likely to be adequately described by a single log-normal distribution (Fig. S8).

#### Directly measuring the ECOFF/ECV from the gWT population

Directly determining the MIC of the 99^th^ percentile from the gWT wild-type population is an attractive option since it requires no further assumptions. This is not usually possible since typically either one cannot discern the wild-type population and/or there are an insufficient number of isolates. The large size of our dataset and the inclusion of genetic information enables us to directly measure the ECOFF/ECV (Fig. 3, S9). Our dataset is enriched for resistance, hence the proportion of resistant samples misclassified as susceptible due to sample mislabelling is likely to be of the order of a few percentage points, even after we have removed some putative mislabelled samples (Methods). This makes directly identifying the 99^th^ percentile challenging, hence we shall also consider the 97.5^th^ and 95^th^ percentiles.

**Figure 3.**
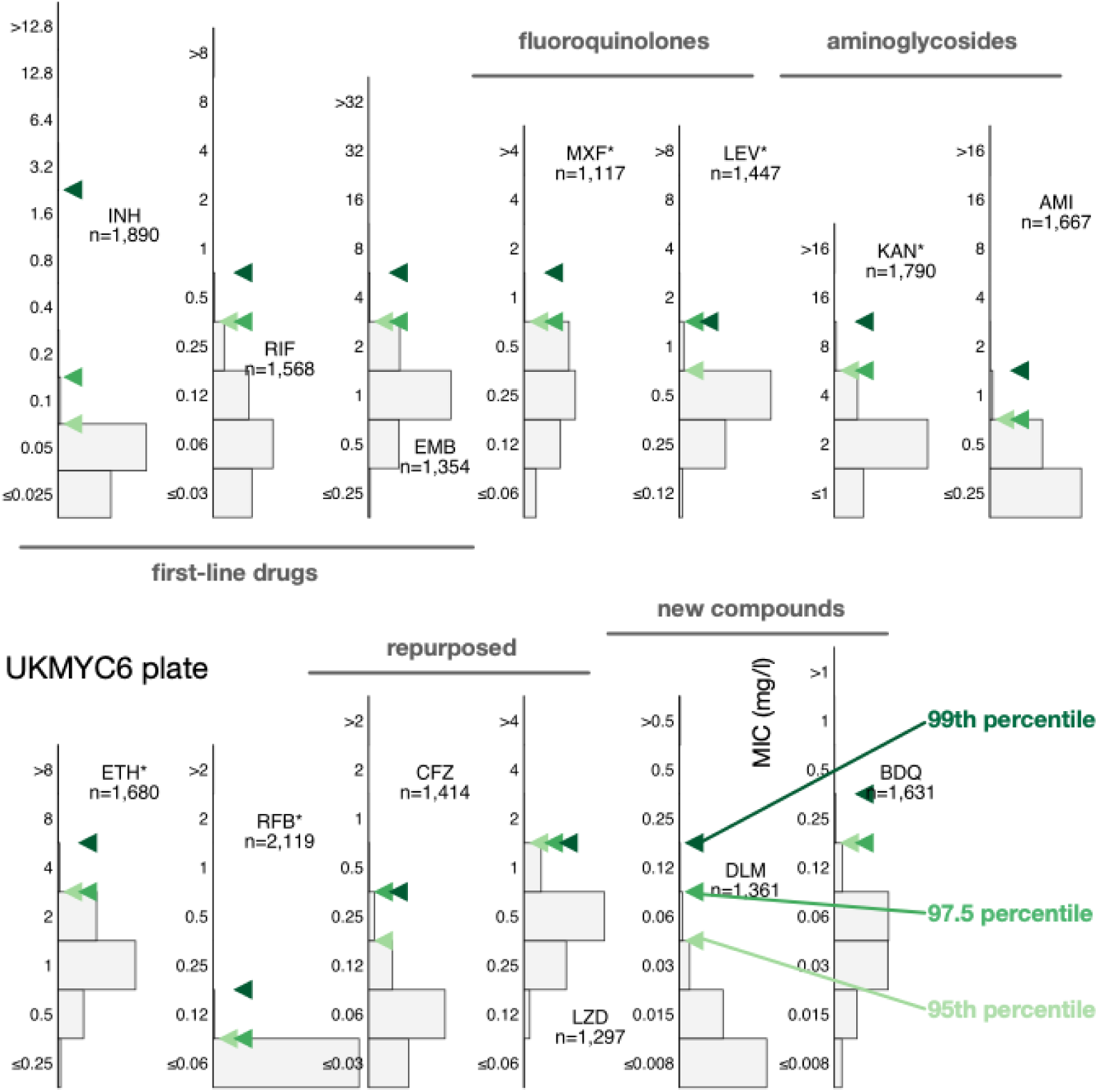
Directly measuring the ECOFF/ECVs from the gWT population on the UKMYC6 plate. To illustrate the sensitivity to the precise percentile used in the definition, the 95th, 97.5th and 99th percentiles are all shown. The analysis and figure can be reproduced (21).

All three percentiles for the MIC histograms of the gWT population are at most two doubling dilutions apart, except for levofloxacin (UKMYC5) and isoniazid (UKMYC6). The latter has an appreciable number of isolates that, despite being classified as gWT, have elevated MICs. These are likely due to some remaining samples that were mislabelled and illustrates the difficulty in using the 99^th^ percentile to define an ECOFF/ECV due to its sensitivity to errors in the dataset, especially when the prevalence of resistance is high, as in the case for isoniazid in our dataset. We shall take forward the values for the 99^th^ percentiles (Table 2) but will bear in mind that a high amount of variation may indicate strain mis-labelling.

#### Interval regression takes account of the truncated distributions

To avoid the identification of the 99^th^ percentile being disproportionately affected by a small number of mislabelled resistant samples one usually fits a log-normal distribution to the pWT (here gWT) population and then calculate from the resulting function the MIC of the 99^th^ percentile. We cannot apply ECOFFinder here since its heuristic requires the presence of non-susceptible isolates in the distribution so we instead simultaneously fit a single log-normal distribution using interval regression to the MIC histograms from both plate designs (Fig. 4, S10, Table 2). With the exceptions of isoniazid, rifabutin and delamanid, the resulting log-normal distributions describe the MIC histograms well, even when there is moderate truncation due to the plate design. The rifabutin MIC distribution is, however, extremely truncated, and hence there are insufficient data to perform a fit - the concentration range for this drug should be lowered in future designs.

**Figure 4.**
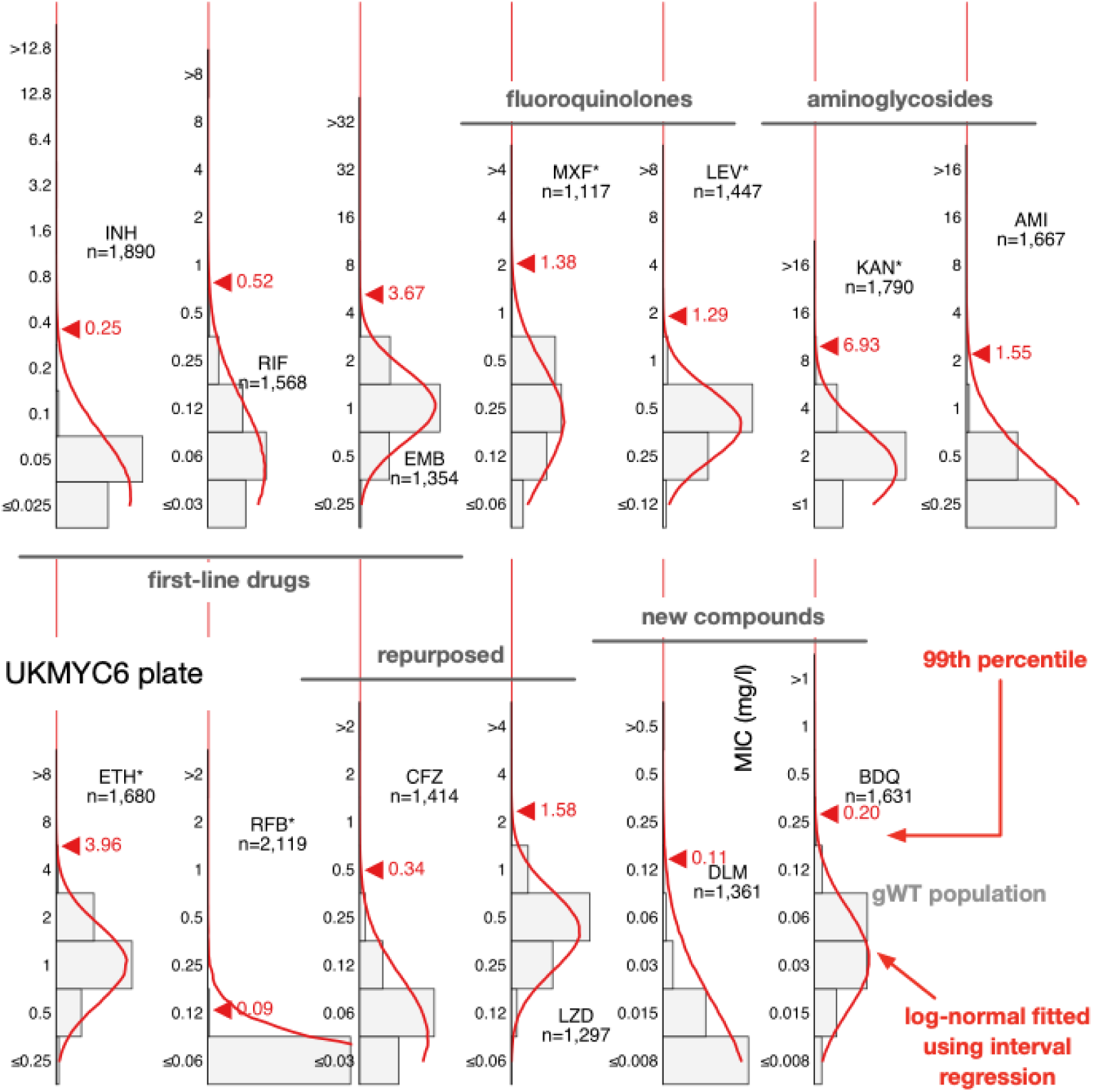
Interval regression is able to fit a log-normal distribution to the MIC histograms of the genotypically wild-type isolates for all 13 drugs on the UKMYC6 plate. Data from both plate designs were considered simultaneously, hence the resulting distributions are those the algorithm considers to best describe both the UKMYC5 (Fig. S10) and UKMYC6 data sets. See the Supplemental Information for the numerical data. The data can be reproduced (21).

#### Proposed ECOFF/ECV values

We infer that direct measurement is the most reliable method since it makes the fewest assumptions. However for drugs where there is variation of more than a doubling dilution between the 95^th^, 97.5^th^ and 99^th^ percentiles, which may indicate the gWT population includes a small but unknown number of isolates with elevated MICs, we shall place greater weight on the result obtained by interval regression. When the MIC histogram is not heavily truncated, we shall also include the 99^th^ percentile reported by ECOFFinder. Note that to convert an MIC that is reported as a real number into an ECOFF/ECV it should be rounded up to the next value in the doubling dilution series. All these data and the resulting ECOFF/ECV values are shown in Fig. 5 & Table 2.

**Figure 5.**
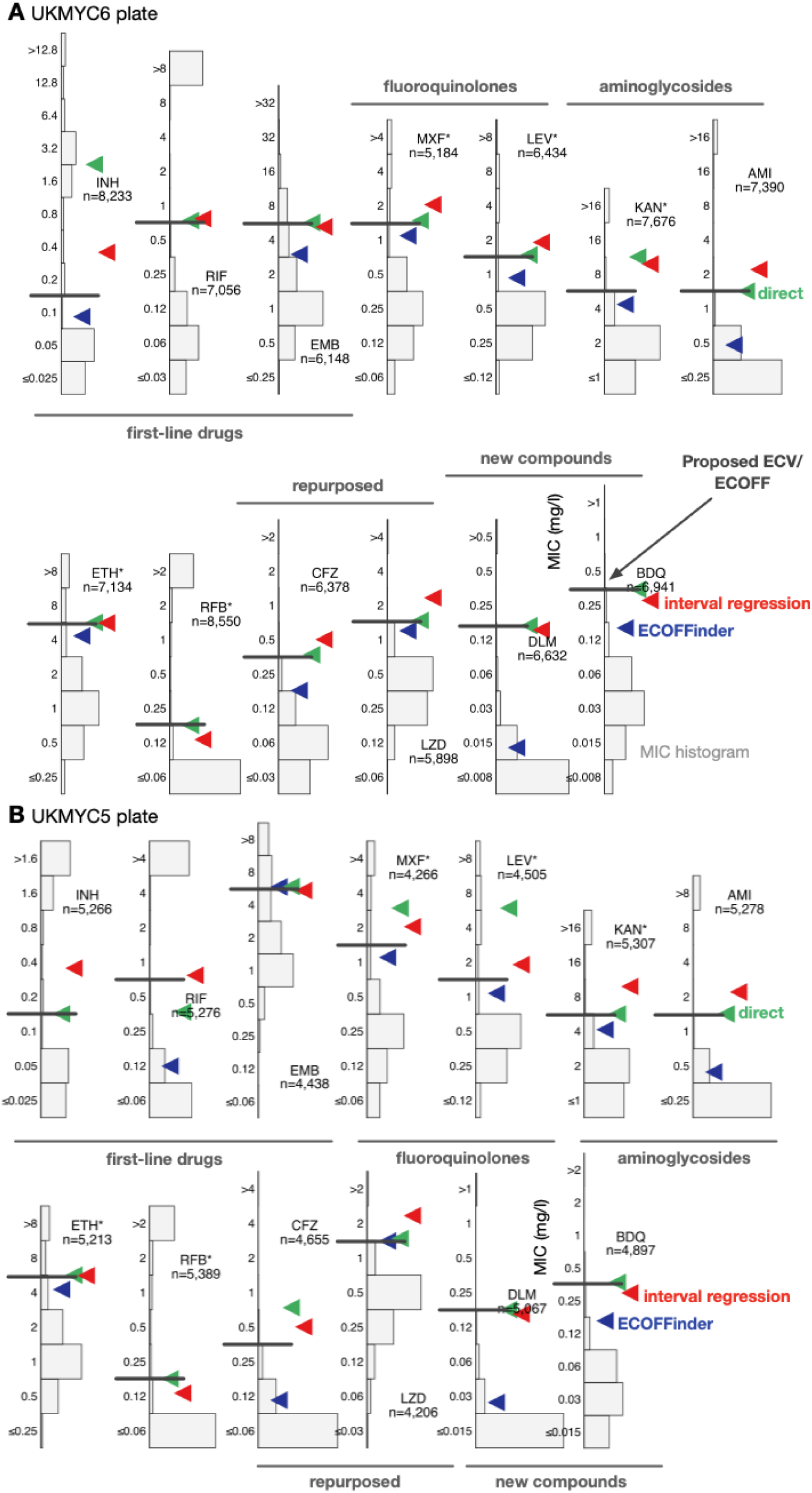
The 99^th^ percentiles of the wild-type populations for the 13 drugs on the (**A**) UKMYC6 and(**B**) UKMYC5 plate designs as calculated by ECOFFinder, direct measurement and interval regression. The ECOFF/ECV values are drawn on each graph as a horizontal line.

The 99^th^ percentile determined by direct measurement for isoniazid for the UKMYC6 dataset was discounted due to the large range of MICs spanned by the percentiles – this is most likely due to resistant samples mis-identified as susceptible due to laboratory mislabelling. INH has the highest prevalence of resistance in the dataset, hence would be affected most. Our starting point is therefore the corresponding value for the UKMYC5 dataset (0.1 mg/L). The ECOFFfinder results were ignored for INH since the gWT MIC histogram is truncated on both plate designs and hence the fits were poor. Visually the gWT MIC histograms (Fig. S8) do not appear to follow a log-normal distribution and consequently interval regression over-estimates the 99^th^ percentile (Fig. 4, S10). The ECOFF/ECV of 0.1 mg/L for Isoniazid is therefore less well supported than the ECOFFs for the remaining drugs.

Direct measurement of the 99^th^ percentiles for rifampicin were 0.5 and 0.25 mg/L for the UKMYC6 and UKMYC5 datasets, respectively. Again, ECOFFinder was not used due to concerns about truncation. Visually the gWT MIC histogram appears more normal in character and the 99^th^ percentile derived from the interval regression fit is 0.52 mg/L. Our proposed consensus ECOFF/ECV for rifampicin is hence 0.5 mg/L. Direct measurement produced a consistent value of 4 mg/L for ethambutol which is supported by interval regression and ECOFFinder for the UKMYC5 dataset (the concentration range on the UKMYC6 plate was more truncated). Both fluoroquinolones behaved similarly: direct measurement gave a value of 1 mg/L for the 99^th^ percentile for both compounds for the UKMYC6 dataset, but 2 mg/L and 4 mg/L for the UKMYC5 dataset (MXF and LEV, respectively). These values for the latter dataset were very sensitive to the exact percentile used in the definition (Fig. S9), again suggesting that these gWT populations may contain a small number of mislabelled resistant samples. Both drugs have the same concentration range on both plate designs, are only moderately truncated and hence ECOFFinder would be expected to give reasonable results. These, along with the result of the interval regression (Fig. 5), result in an ECOFF/ECV of 1 mg/L for both fluoroquinolones.

Direct measurement indicates the 99^th^ percentile for kanamycin is 8 mg/L and 4 mg/L for the UKMYC6 and UKMYC5 datasets, respectively, whilst it produces the consistent value of 1 mg/L for amikacin. The MIC histogram of the latter is too truncated for ECOFFinder to function correctly and visually the log-normal fitted by interval regression appears to have over-estimated the 99^th^ percentile as 1.6 mg/L, hence we propose and ECOFF/ECV for amikacin of 1 mg/L. The kanamycin MIC histograms are less truncated and interval regression better describes the gWT population; these data support an ECOFF/ECV of 4 mg/L for kanamycin. For ethionamide direct measurement produces a consistent value of 4 mg/L which is supported by interval regression and ECOFFinder. The MIC histogram of rifabutin is extremely truncated and hence only direct measurement is likely to be effective; it estimates that 0.12 mg/L to be the 99^th^ percentile for both datasets, which is therefore our ECOFF/ECV.

Direct measurement yields consistent values of 0.25 mg/L and 1 mg/L for bedaquline and linezolid respectively, with each supported by both interval regression and ECOFFinder. Our ECOFF/ECV for delamanid is 0.12 mg/L since this is the direct measurement which is the same for both datasets and it is supported by interval regression. Lastly, direct measurement for clofazimine suggests the 99^th^ percentiles are 0.25 mg/L and 0.5 mg/L for the UKMYC6 and UKMYC5 datasets, respectively. The latter has more variation and hence we propose its ECOFF/ECV is 0.25 mg/L.

##### Comparison against genetic variants known to confer resistance

Using the subset of the isolates with genetic information, we can examine our proposed ECOFF/ECVs by plotting the MIC histograms of several genetic variants that are widely accepted to confer resistance to key anti-TB drugs (Fig. 6 & S11). The *rpoB* S450L and *katG* S315T single nucleotide polymorphisms substantially increase the MICs of rifampicin and isoniazid, respectively, and the majority (96.9% & 99.5%) of isolates with these mutations had MICs greater than the ECOFF/ECV. The c-15t mutation in the promoter of the *fabG1/inhA* operon was associated with borderline (0.2 mg/L) isoniazid MICs unless present in combination with a *katG* S315T mutation (MIC >1.6 mg/L), as observed elsewhere (22, 28). It is likely that this promoter mutation, and others like it, are responsible for the small peak in the MIC histogram observed for isoniazid at 0.2 mg/L.

**Figure 6.**
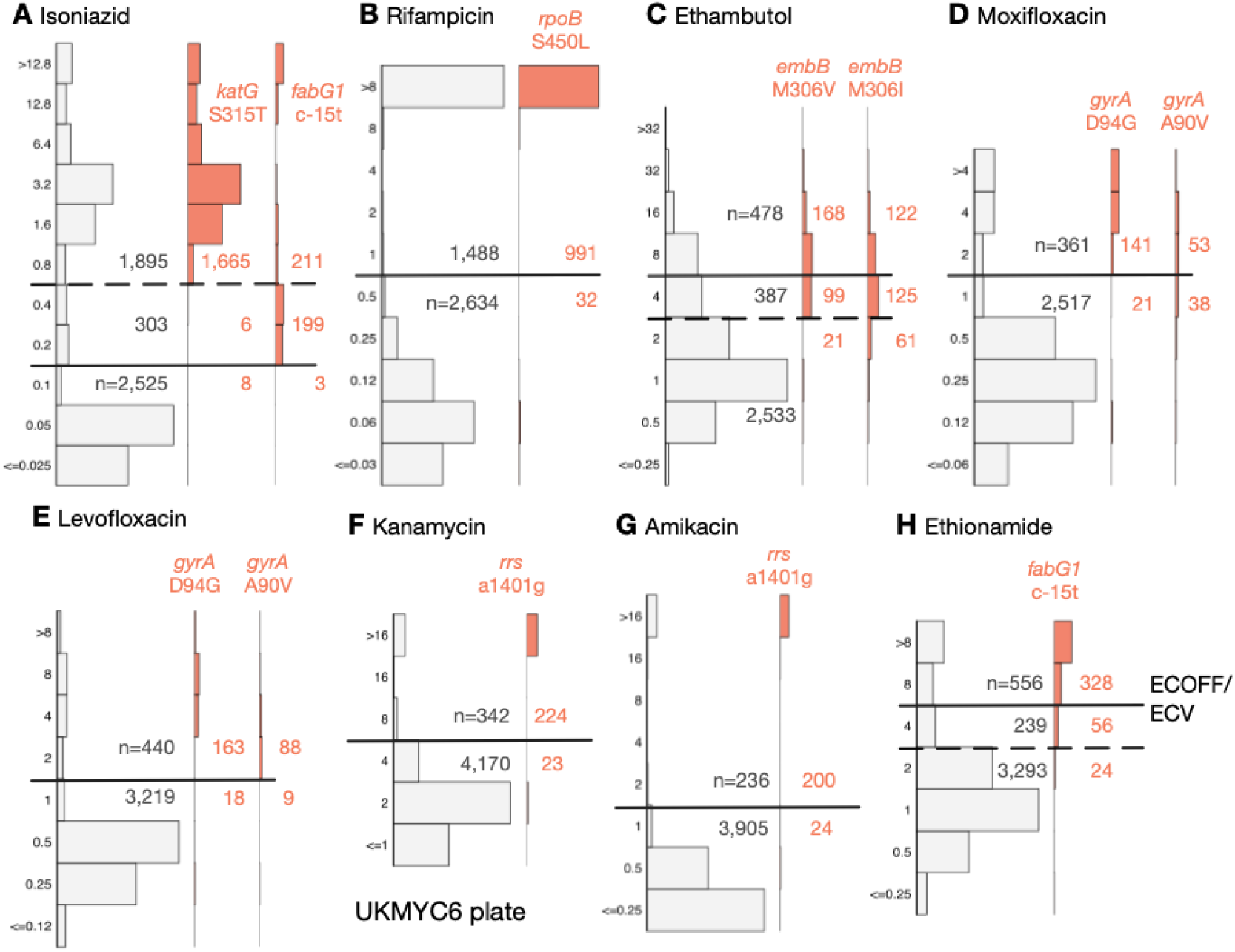
The MICs of isolates containing genetic variants known to confer resistance to different drugs tend to lie above the ECOFF/ECV on the UKMYC6 plate. The number of isolates lying above and below the ECOFF/ECV is annotated. The dashed line indicates the margin of a proposed ‘borderline’ category for isoniazid, ethambutol and ethionamide. The same analysis has been repeated on the UKMYC5 dataset (Fig. S11) and can be reproduced (21).

Substituting isoleucine or valine at position 306 in the *embB* gene was associated with elevated ethambutol MICs, however, the increase in MIC is much less than observed for either of the rifampicin or isoniazid resistance-conferring mutations mentioned above, leading to only 58.3% and 39.6% of isolates containing these mutations, respectively, having an MIC above the ECOFF/ECV. This is expected since it is known that isolates containing these variants can have variable or discordant MGIT results (29). For both fluoroquinolones, the *gyrA* D94G mutation increases the MIC more than the *gyrA* A90V mutation (30), however for levofloxacin the wild-type and non wild-type populations appear slightly better separated with the result that for these mutations 90.0% & 90.7% of isolates lie above the ECOFF/ECV whilst for moxifloxacin the equivalent values are 87.0% & 58.2%. The majority of isolates (90.7% & 89.3%) with the a1401g mutation in the *rrs* gene have an MIC above the ECOFF/ECV for kanamycin and amikacin, respectively. Finally, whilst the c-15t mutation in the promoter of the *fabG1/inhA* operon increases the MIC of ethionamide more than it does isoniazid, only 80.4% of isolates with this variant lie above the ECOFF/ECV.

##### A role for a Borderline category?

The ECOFF/ECV merely defines an MIC below which the majority of the ‘wild-type’ isolates should lie. It does not necessarily follow that the majority of non wild-type isolates have an MIC above the ECOFF/ECV and therefore care needs to be taken when using an ECOFF/ECV to define susceptibility and resistance. Although for most drugs an MIC below or equal to the ECOFF/ECV can be categorised as ‘susceptible’ and those with MICs above the ECOFF/ECV are ‘resistant’, the MIC histograms of isoniazid, ethambutol and ethionamide are more complex. There is genetic evidence (Fig. 6) that this is due to a multitude of genetic variants, each with a different effect on the MIC. We therefore propose a third category, ‘borderline’, for isoniazid, ethambutol and ethionamide (Fig. 6, Table 3).

**Table 3.** The proposed ECOFF/ECV values and suggested borderline MICs for three compounds.

| Drug | ECOFF/ECV | Borderline |
| --- | --- | --- |

|  |  | (mg/L) | (mg/L) |
| --- | --- | --- | --- |
| Isoniazid | INH | 0.1 | 0.2, 0.4 |
| Rifampicin | RIF | 0.5 | - |
| Ethambutol | EMB | 4 | 4 |
| Moxifloxacin | MXF | 1 | - |
| Levofloxacin | LEV | 1 | - |
| Kanamycin | KAN | 4 | - |
| Amikacin | AMI | 1 | - |
| Ethionamide | ETH | 4 | 4 |
| Rifabutin | RFB | 0.12 | - |
| Clofazimine | CFZ | 0.25 | - |
| Linezolid | LZD | 1 | - |
| Delamanid | DLM | 0.12 | - |
| Bedaquiline | BDQ | 0.25 | - |

##### Validation by comparison to MGIT and MODS results

The resistance of a subset of isolates was independently tested to a range of compounds using either the Mycobacteria Growth Indicator Tube (MGIT) system or the microscopic-observation drug-susceptibility (MODS) assay (31). We can therefore validate our MIC-based categorisation by directly comparing between the binary (or ternary) phenotype derived from an MIC and the result from one of these well-established clinical microbiology methods (Fig. 7 & S12, Table S10).

**Figure 7.**
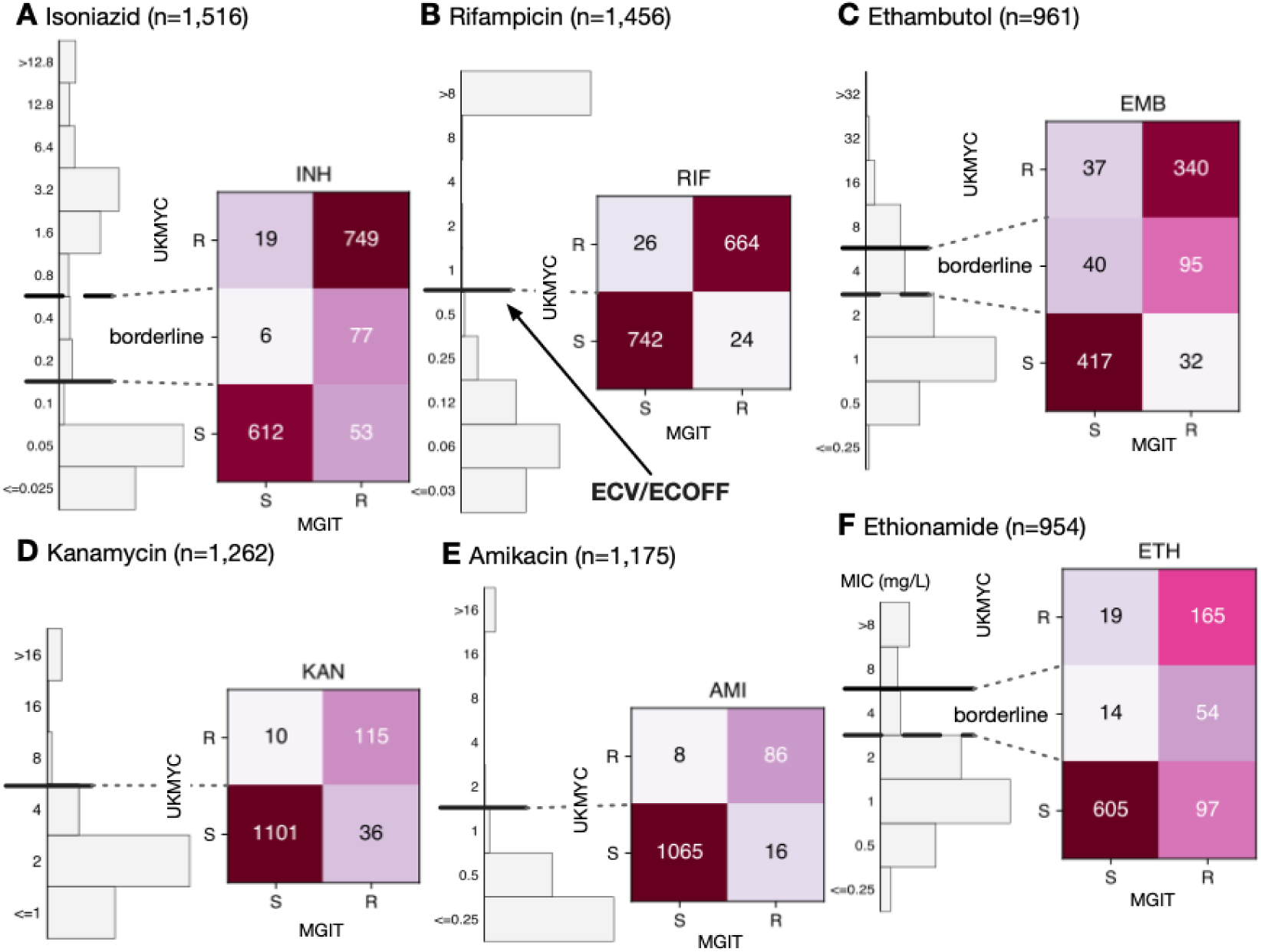
The binary (or ternary) classification derived from the MIC using the ECOFF/ECVs and MIC-based categorisation in Table 3 agrees well with MGIT results for the samples for (**A**) isoniazid, (**B**) rifampicin, (**C**) ethambutol, (**D**) kanamycin, (**E**) amkacin and (**F**) ethionamide. These data and figures can be reproduced (21).

The agreement between MGIT and UKMYC is good, with a sensitivity of 93.4% and a specificity of 97.0% (Table S10). Since the borderline category lies above the ECOFF/ECV, it is interpreted as providing a way of discriminating between isolates with a moderately elevated MIC and those with a high MIC. For rifampicin, the agreement between MGIT and UKMYC is excellent with a sensitivity of 96.5% and a specificity of 96.6%. The borderline category for ethambutol provides a “buffer zone” since isolates with an MIC of 4 mg/L are only 70.4% resistant according to MGIT. Ignoring these isolates, the sensitivities and specificities are 91.4% and 91.9%, respectively, for ethambutol. Since the ‘borderline’ category in this case lies below the ECOFF/ECV, these isolates would otherwise be classified as ‘susceptible’ but in reality have a mixed character.

Hence not assigning a borderline category would result in the sensitivities and specificities becoming 72.8% and 92.5%, respectively.

The aminoglycosides behave similarly to one another with sensitivities and specificities of 76.2% and 99.1% for kanamycin and 84.3% and 99.3% for amikacin, respectively. The ‘borderline’ category for ethionamide (MIC of 4 mg/L) are 79.4% resistant according to MGIT. Excluding these isolates, the sensitivity is 63.0% and 97.0%, respectively. Limited number of isolates were tested for moxifloxacin or levofloxacin resistance using MGIT (Fig. S12). Although large number of isolates were tested for clofazimine and linezolid resistance by MGIT (Fig. S12), the low prevalence of resistance ensures no useful conclusions can be drawn.

A different set of samples were tested in parallel using the MODS assay (Fig. S13, Table S11). The sensitivities and specificities for isoniazid (n=1,888) and rifampicin (n=1,857) were 95.3% & 98.9% and 95.1% & 99.2%, respectively.

## DISCUSSION

We have proposed epidemiological cut-offs (ECOFF/ECVs) for research-use for 13 different anti-TB compounds for the UKMYC series of broth microdilution plates using an aggregated dataset of 20,637 tuberculosis samples collected worldwide by 14 CRyPTIC laboratories based in 11 countries on five continents. The UKMYC6 plate design (Fig. 1B) not only contain the first-line drugs rifampicin, isoniazid and ethambutol, but also all of the Group A drugs, one of the two Group B compounds (clofazimine) and five of the seven Group C medicines recommended by the World Health Organisation for treating cases of multi-drug resistant tuberculosis (19). As such these plates offer near comprehensive phenotypic AST as well as a standardised, scalable phenotype that, once analysed with linked genomic data, will inform clinical decisions where routine diagnostics switch to genome sequencing.

We caution that whilst the ECOFF/ECVs proposed herein have been derived using the largest collection of *M. tuberculosis* samples to date, the methods do not conform with those laid out by EUCAST. That said, our analyses illustrate that the EUCAST definition of an ECOFF/ECV as the 99^th^ percentile of the wild-type population (20) is difficult to apply in practice since firstly it is not always possible to define which isolates are *phenotypically* wild-type without engaging in a circular argument. We were able to avoid this here by defining a *genotypically* wild-type (gWT) population. The second problem is that using the 99^th^ percentile to define the ECOFF/ECV places a very stringent upper limit on the total error rate which becomes harder to meet as the prevalence of resistance in any dataset increases. Despite our efforts, we see evidence that our gWT populations for some compounds contain >1% resistant isolates for some drugs which e.g. hampers the use of direct measurement. In contrast the CLSI have a less-stringent definition for the ECOFF/ECV which avoids this issue (32) but in turn can create inconsistencies between studies. As suggested elsewhere, using a lower percentile (e.g. 97.5^th^) could help (22).

The CLSI recently proposed breakpoints for the MYCOTB plate (33). There are a few minor differences: our proposed ECOFF/ECV for rifampicin is one doubling dilution lower at 0.5 mg/L. The impact of this is difficult to assess due to the paucity of isolates with MICs of 0.5 and 1.0 mg/L. For isoniazid, although it is not possible to make an exact comparison between the ECOFF/ECVs since the doubling dilution series used on the UKMYC plates and by the CLSI are different, the value proposed by CLSI (0.12 mg/L) is close the value proposed here (0.1 mg/L). The CLSI breakpoints for ethambutol exactly agree with the MIC-based classification adopted by CRyPTIC. Our ECOFF/ECVs are different to those of a recent MYCOTB study (22), however we note that the number of samples was modest (385) and originated from a single country. In addition, ECOFF/ECVs were determined using ECOFFinder, which given the truncated nature of the MIC histograms for many of the drugs, is not now advised and may have biased some of the results. Our ECOFF/ECVs for rifampicin and isoniazid are, however, consistent with a recent recommendation made by the WHO, albeit for MGIT

Critical concentrations for several of the drugs on the UKMYC plates (which are inoculated with 7H9 growth media) exist for *M. tuberculosis* grown in other growth media, such as Löwenstein-Jensen, 7H10 and 7H11, and also other AST methods, such as the BACTEC Mycobacterial Growth Indicator Tube 960 (34–36). Caution must, of course, be applied when comparing cut-offs derived using fundamentally different growth media and AST methods.

The ECOFF/ECVs for nine drugs proposed by a series of 7H10 agar studies all either agree or are one doubling dilution different to our proposed ECOFF/ECVs for the UKMYC plates (37–39). More recently, there has been a push to set breakpoints for the new compounds delamanid and bedqauline so that AST can be performed for these important drugs (40). An early MGIT study using 194 isolates proposed an ECOFF/ECV for delamanid of 0.125 mg/L (41), which is identical to our value. An ECOFF/ECV of 0.125 mg/L for bedaquiline on broth microdilution plates was proposed (42), however the 95^th^ percentile of the wild-type population was used to define the ECOFF/ECV since CLSI guidelines were followed (32) and ECOFFinder was used despite the truncated nature of the MIC histograms. This value was supported by two subsequent studies, the first of which showed that that the sensitivity and specificity is maximised with a ECOFF/ECV of 0.12 mg/L compared to 0.25 mg/L (43). The second confirmed this value, however also stated that the 99^th^ percentile of the wild-type population was 0.25 mg/L (44). This illustrates that the exact value can be difficult to pin down when different ECOFF/ECV definitions are used; hopefully the number and diversity of samples in our study will help resolve this important question. Lastly, the ECOFF/ECVs proposed here lie within the range of breakpoints recommended by the WHO for different growth media, with the exception of clofazimine for which the WHO recommends a cut-off of 1 mg/L in MGIT (35, 36).

Deriving ECOFF/ECVs from MICs relies on several assumptions, foremost that applying a binary resistant/susceptible classification to a clinical infection is a reasonable and helpful way to proceed. That simplifying the description of the results of clinical microbiology investigations helps interpretation is not in doubt (45), however problems with reproducibility can arise depending on the character of the underlying MIC histogram. If the MIC histogram is ‘bimodal’ (i.e. has two narrow peaks separated by an interval greater than their individual variance) then placing the ECOFF/ECV between the peaks leads to a helpful and reproducible classification system (46). On the UKMYC series of plates, the only drugs that conform to this ideal are the rifamycins and the aminoglycosides; the other compounds either have more complex distributions (INH, EMB, MXF, LEV, ETH) or resistance is not sufficiently prevalent for us to fully characterise their MIC distributions (CFZ, LZD, DLM, BDQ).

There is a further implicit (weak) assumption that the ‘susceptible’ and ‘resistant’ subpopulations can each be described by a *single* MIC distribution, which is not necessarily true, as exemplified by the effect of the *fabG1* promoter mutations on isonizaid (Fig. 6A) (28). In addition this assumption implies that different lineages behave similarly when exposed to an antibiotic, which is unlikely to be true (47). Finally, the wild-type distribution is usually assumed to be log-normal, however our data do not support this for all drugs (e.g. LEV, Fig 4) resulting in the log-normal distributions fitted by interval regression apparently over-estimating the 99^th^ percentile. Should this turn out to be generally true, this would invalidate methods based on fitting such distributions, making direct measurement more appealing (25).

One can deconstruct the error in determining an ECOFF/ECV using a microtitre plate into sample selection biases, data entry and labelling errors, inoculation and incubation error, measurement error, error in defining the wild-type population, uncertainties arising from censored data and error in fitting a curve to the resulting MIC histogram. In addition to the obvious benefits in collecting such a large and diverse dataset (Table 1, Fig 1, Table S2), we have been careful to minimize measurement error (Fig. S3) and have also used a principled method to attempt to remove some putative mislabelled samples (Methods). By defining a genotypically wild-type population and either applying interval regression to fit normal distributions (Fig. 4) or directly measuring the 99^th^ percentile (Fig. 5), we have also minimized the final two sources of error. Despite these steps, further sources of error no doubt remain. Another key weakness of this study is the lack of pyrazinamide, which due to its preference for acidic conditions, is currently unable to be successfully incorporated onto broth microdilution plates, although there is hope that this could be rectified in future (48).

The debate about how to define and calculate ECOFF/ECVs will continue and new approaches will be suggested (49–52). However it evolves, larger and more geographically diverse tuberculosis datasets, such as presented here, will bring more confidence and rigour to the antibiotic susceptibility testing of clinical tuberculosis samples. We hope also, that as clinical microbiology transitions into a data-driven science, our proposed method of directly measuring the required percentile of the gWT population will gain traction due to its simplicity and reproducibility as the genetics of *M. tuberculosis* resistance becomes better understood and accepted.

Although the main objective of the CRyPTIC project is to map the genetic variations in the *M. tuberculosis* genome that confer resistance to many antibiotics, the sheer number of samples collected provides a body of evidence to support the use of 7H9 broth microdilution plates in clinical mycobacteriology. Applications potentially include antibiotic susceptibility testing for samples that are predicted to be MDR or XDR by the GeneXpert RIF/MDR assay system or surveying the prevalence of different patterns of resistance by region or country, allowing regional regimens to be designed and their impact monitored. Finally, even in settings which adopt genetics-based clinical microbiology (11), it would be prudent to maintain culture-based testing not only to identify new genetic variants as they arise but also to continuously monitor the performance of the genetic resistance catalogue which are likely to change over time as such catalogues are only likely partly causal.

In future work the CRyPTIC project will apply the ECOFF/ECVs proposed here not only to further optimise a genetic catalogue for the first-line anti-TB compounds (6) but also to extend coverage to second-line, repurposed and new compounds, with the aim of covering as many of the drugs recommended by the WHO for treating MDR and XDR tuberculosis (19). Clearly the numerical data being collected by the consortium also lends itself to the development of a genetic catalogue for anti-TB compounds that can make *quantitative* predictions; such a catalogue would naturally take account of additivity, epistatis and non-linear effects. Finally, the tools and data-driven approaches developed here could be applied to other pathogens, especially other mycobacteria.

## METHODS

### Ethics review

Approval for the CRyPTIC study was obtained by Taiwan Centers for Disease Control IRB No. 106209, University of KwaZulu Natal Biomedical Research Ethics Committee (UKZN BREC) (reference BE022/13), University of Liverpool Central University Research Ethics Committees (reference 2286), Institutional Research Ethics Committee (IREC) of The Foundation for Medical Research, Mumbai (Ref nos. FMR/IEC/TB/01a/2015 and FMR/IEC/TB/01b/2015), Institutional Review Board of P.D. Hinduja Hospital and Medical Research Centre, Mumbai (Ref no. 915-15-CR [MRC]), scientific committee of the Adolfo Lutz Institute (CTC-IAL 47-J / 2017) and in the Ethics Committee (CAAE: 81452517.1.0000.0059) and Ethics Committee review by Universidad Peruana Cayetano Heredia (Lima, Peru) and LSHTM (London, UK). No ethics approval was required for the remaining laboratories since at no time was any patient identifiable information shared with the consortium.

### Sample selection

The CRyPTIC project aimed for around half the samples collected to be susceptible to the first-line compounds with the remainder MDR/XDR. There was, however, large variation between the different participating laboratories.

### Incubation and inoculation protocol

Each laboratory followed a standard operating protocol laid out by the CRyPTIC consortium, which was similar to that described previously (17). Clinical samples were sub-cultured either using Lowenstein-Jensen tubes, 7H10 agar plates or MGIT tubes. The protocol specified that first a suspension at 0.5 McFarland standard in saline Tween with glass beads (Thermo Fisher, Scientific Inc., USA) from 20-to 25-day-old colonies. These were then diluted 100-fold by adding 100 μl of suspension to 10 ml of enriched 7H9 broth (17). A semi-automated Sensititre Autoinoculator (Thermo Fisher, Scientific Inc., USA) was used to dispense 100 μl of inoculum (1.5 × 10^5^ CFU/ml, with approximate range from 5 × 10^4^ CFU/ml to 5 × 10^5^ CFU/ml) into a well of a UKMYC5/6 microdilution plate. The plate was then sealed using transparent plastic provided by the manufacturer. The UKMYC5 and UKMYC6 microdilution plates were designed by the CRyPTIC consortium and manufactured by Thermo Fisher Inc., U.K. The drugs included and their concentrations are described in Fig. 1. Delamanid and bedaquiline pure substances were provided by Otsuka Pharmaceutical Co., Ltd. and Jannsen Pharmaceutica, respectively. The H37Rv ATCC 27294 was used to perform periodic quality control runs since it is susceptible to all the drugs on both plate designs.

### Measurement of MICs after 14 days incubation

In each laboratory a scientist read each plate after 14 days incubation using a Thermo Fisher Sensititre Vizion digital MIC viewing system, with results entered via a bespoke web portal (https://clires2.oucru.org). In those cases where this was not possible, spreadsheets were sent. A photograph was also taken using the Vizion system and also stored in CliRes2. Two laboratories used a mirrored-box to read the plates and one of these also took a photograph using a DSLR. A plate was marked as invalid if it did not have adequate bacterial growth in both positive control wells. A small subset of plates with poor growth at day 14 were incubated for a further week and then read again.

### Other AST measurements

Where available, the results of standard AST tests conducted by the participating laboratory were also entered via the CliRes2 online portal, or in some cases shared via spreadsheet. The methods used were mainly either the Mycobacteria Growth Indicator Tube (MGIT) system or the microscopic-observation drug-susceptibility (MODS) assay (31). All MGIT tests used standard critical concentrations (CC) – for the moxifloxacin results only those with a CC of 0.5 mg/L were included.

### Genetic sequencing and interpretation

Sequencing arrangements differed slightly between each CRyPTIC participating laboratory. All sequencing was performed using Illumina machines and hence the input to our genetic sequencing pipelines was a matched pair of FASTQ files containing the short reads. Data integrity was ensured throughout by tracking the MD5SUM hashs of the FASTQ files.

Human and HIV reads were removed from the raw sequence data as follows. Reads were mapped to the reference genome H37Rv, the human genome version GRC38, the HIV reference NC_001802.1, various other viral genomes (so that, if any reads mapped to HIV, no-one would only know that they mapped to some virus), and nasopharyngeal flora genomes from the human microbiome project, using BWA MEM. First, a read pair was kept if either read matched H37Rv, then removed if either read matched one of the other genomes, and finally kept if both reads were unmapped.

Variants were initially called using SAMtools and Cortex, two variant callers with orthogonal strengths (samtools a high sensitivity SNP caller, and cortex a high specificity SNP and indel caller). These calls were then passed to the adjudication software minos, which produces a graph representation of the reference genome plus conflicting calls from the two callsets, and then remaps reads to the graph to adjudicate statistically. This adjudication process, and the performance of the combined samtools/cortex callset, are documented (53). All of this process, including versions of samtools and cortex and the reference genomes for filtering) is encapsulated in Clockwork version 0.8.3 (54).

Samples were excluded from the dataset if they had either more than 100,000 unfiltered samtools variant calls (a weak filter applied to detect samples contaminated with the wrong species) or an average read coverage of 15 or less when mapped to reads covering the H37Rv reference. The samples that pass these criteria and have paired phenotype data are named the GPI (geno-pheno intersection). Variant calls were removed if they overlapped a set of masked positions as previously defined (55). This mask consists of 324,971 positions from the H37Rv reference with self-blast matches, and can be found here: https://github.com/iqbal-lab-org/cryptic_tb_callable_mask/commit/43ec21319209b23f648f32e4868bdf07cf09f2a0.

Version 3 of the H37Rv strain (NC_000962.3) was used as the TB reference genome throughout. The resulting VCF files were then transferred to the CRyPTIC data warehouse where they were interpreted.

### Genetic resistance catalogue

A hybrid TB genetic resistance catalogue was constructed by merging two published catalogues, the first more recent catalogue contained rows for the four first-line drugs (INH, RIF, EMB, PZA) (6). The second also contained rows for MXF, LEV, STM, OFX, AMI, KAN, CAP, ETH, LZD, CFZ, DLM, BDQ, RFB, PTO, PAS (8). Since each catalogue was constructed with respect to version 2 of the H37Rv *M. tuberculosis* reference genome, they were first translated to version 3 of the reference. These catalogues are freely available to download (56) and use a standard grammar, GARC, that is both machine- and human-readable. To avoid putting as few assumptions into downstream code as possible, default rules are included that e.g. specify that non-synonymous amino acid mutations that match no other row have an unknown effect. The hybrid catalogue was constructed by taking the rows for the first-line compounds from the first catalogue and rows for all other drugs from the second. This catalogue, called CRyPTICv1.31, is freely available for download (56) and is also provided in the attendant repository (21).

### Genetic analysis

Each sample VCF was compared to a reference genome object using the Python gumpy module (57), thereby creating a table of genetic variants (both single nucleotide polymorphisms, SNPs, and insertions/deletions). Both the individual SNPs were stored and also their aggregated effect on any coding region of gene encoding a protein sequence. An intergenic region of up to 100 bases upstream of the start codon was assumed to be the promoter sequence and hence was associated with the gene. This list of variants was then parsed by a second bespoke Python module, piezo, that reads the hybrid catalogue and understands the GARC grammar and so returns a resistant, susceptible or unknown prediction for each drug in the catalogue (58). The species and, if *M. tuberculosis*, lineage of all samples was determined by SNP-IT (23).

### Data warehousing

With the exception of the compressed FASTQ files, all data (VCFs, images, MIC metadata, genetic variants and catalogue predictions) were aggregated and stored in a hierarchical file system using the Python datreant 1.0.2 module (59) which allowed for data discovery, tagging and filtering. Updates were performed by inhouse Python scripts. Plate metadata was downloaded from CliRes2 using the zeep Python SOAP client.

### Quality assurance of minimum inhibitory concentration readings

Central to our quality assurance (QA) process is the photograph taken of the plate after 14 days of incubation using the Vizion instrument by the laboratory scientist. Images were deduplicated by checking the MD5SUM was unique. The remaining images were first read by bespoke software, AMyGDA (60, 61), which detects the locations of the wells and, by measuring the growth in each well, estimates an MIC for all drugs. For 54.7% of all measurements the MICs measured by the laboratory scientist and AMyGDA were identical (Fig. S4, S5A) and therefore passed the quality control process.

Images of the 45.3% of cases where these two methods disagreed were uploaded to a Citizen Science project, hosted by the Zooniverse platform, called BashTheBug (62). Each image was classified by at least 11 different volunteers and the median reading was taken to be the consensus. In 38.1% of the images sent (17.3% of the total) the consensus MIC agreed with the MIC measured by the laboratory scientist using the Vizion instrument. Visual inspection of a random subset (Fig. S5B) suggested that these were mostly cases where AMyGDA incorrectly estimated the MIC, usually because the growth was too small to be programmatically detected. For a smaller proportion (12.0% of the images completed by BashTheBug, 5.4% of the total – Fig. S3), the BashTheBug consensus agreed with the MIC measured by AMyGDA. Visual inspection of a random subset (Fig. S5C) indicated that, for the most part, these were errors made by the laboratory scientist. An error rate of 5.4% for a subjective laboratory-based measurement is reasonable and catching and correcting these errors is the main goal of this quality control process. Overall, therefore, we have a high degree of confidence in 77.4% of the MIC measurements since two or more independent methods concur on the value. Finally, in 22.6% of cases all three methods gave a different answer (Fig. S5D); these are excluded from further analysis. All these proportions are averaged over all drugs; there is significant variation between drugs (Table S7 & S8).

### Putative mislabelled samples

Some 44 samples were assumed to be mislabelled samples as defined by being genotypically wild-type but having both an INH MIC ≥ 1.6 mg/L and an RIF MIC ≥ 4 mg/L. This corresponds to 0.8% of the dataset which is likely an underestimate. All 44 samples were removed.

### ECOFFinder

A version of ECOFFinder (ECOFFinderXL2011forMac.xlxs) that worked on Microsoft Excel running on Apple Mac computers was provided by Dr Claudio Köser (25).

### Interval regression

The intreg function in STATA version v15.1 (Stata Corp.) was used.

### Data analysis and graphs

All data analysis, with the exception of the interval regressions and ECOFFinder, were performed using Python 3.8 in conjunction with Pandas 1.2.1 (63), numpy 1.19.5 (64). Graphs were plotted using matplotlib 3.3.4 (65) and GeoPandas 0.8.2.

### Reproducibility

The raw data (photographs of 96-well plates, genetic variant call files) along with a series of data tables related by a schema can be downloaded from the European Bioinformatics Institute at http://ftp.ebi.ac.uk/pub/databases/cryptic/. In addition, one can reproduce nearly all the tables and figures, along with the Supplemental Data, in a browser window (i.e. no installation required) using Python code we have made publicly available (21).

## Data Availability

The numerical data used to plot Figures 2 and 3 from which the ECOFF/ECVs are derived is included in a Supplemental CSV file.
In addition, one can reproduce nearly all the tables and figures, along with the Supplemental Data, in a browser window (i.e. no installation required) using Python code we have made publicly available here: https://github.com/fowler-lab/cryptic-ecoffs

## ACKNOWLEDGEMENTS

We are grateful to Claudio Köser for providing the ECOFFinder programme and for helpful comments, the EUCAST ESGMYC subcommittee chaired by Emmanuelle Cambau, especially John Turnidge, for helpful discussions and all the BashTheBug volunteers for the time and energy they have contributed. We thank Faisal Masood Khanzada and Alamdar Hussain Rizvi (NTRL, Islamabad, Pakistan), Angela Starks and James Posey (Centers for Disease Control and Prevention, Atlanta, USA), and Juan Carlos Toro and Solomon Ghebremichael (Public Health Agency of Sweden, Solna, Sweden).

## AUTHOR CONTRIBUTIONS

- All contributed laboratories collected samples and provided data.
- DWC, TEAP, SH, ALGC, AWS, TMW, PWF, DMC designed the study.
- PWF, SH, ALGC and retrieved and analysed the MIC data.
- ZI, MH, JK and PWF analysed all the genetic information.
- PWF, ASW, TMW performed all analysis.
- PWF wrote the manuscript with all partners offering feedback.

## REFERENCES

1. World Health Organization (2020) Global Tuberculosis Report.

2. Enkirch T, et al. (2020) Systematic Review of Whole-Genome Sequencing Data To Predict Phenotypic Drug Resistance and Susceptibility in Swedish Mycobacterium tuberculosis Isolates, 2016 to 2018. Antimicrob Agent Chemo 64(5):2–5.

3. Gygli SM, et al. (2019) Whole-Genome Sequencing for Drug Resistance Profile Prediction in Mycobacterium tuberculosis. Antimicrob Agents Chemother 63(4):1–13.

4. Ransom EM, Potter RF, Dantas G, Burnham C-AD (2020) Genomic Prediction of Antimicrobial Resistance: Ready or Not, Here It Comes! Clin Chem 66(10):1278–1289.

5. Phelan J, et al. (2016) The variability and reproducibility of whole genome sequencing technology for detecting resistance to anti-tuberculous drugs. Genome Med 8(1):1–9.

6. The CRyPTIC Consortium, 100000 Genomes Project (2018) Prediction of Susceptibility to First-Line Tuberculosis Drugs by DNA Sequencing. New Eng J Med 379(15):1403–1415.

7. Walker TM, et al. (2015) Whole-genome sequencing for prediction of Mycobacterium tuberculosis drug susceptibility and resistance: a retrospective cohort study. Lancet Infec Dis 15(10):1193–202.

8. Miotto P, et al. (2017) A standardised method for interpreting the association between mutations and phenotypic drug resistance in Mycobacterium tuberculosis. Eur Respir J 50(6):1701354.

9. World Health Organization (2021) Catalogue of mutations in Mycobacterium tuberculosis complex and their association with drug resistance Available at: https://www.who.int/publications/i/item/9789240028173.

10. Walker TM, et al. (2021) The 2021 WHO Catalogue of Mycobacterium Tuberculosis Complex Mutations Associated with Drug Resistance: A New Global Standard for Molecular Diagnostics. SSRN Electron J 09(5):7352–7363.

11. Walker TM, et al. (2017) Tuberculosis is changing. Lancet Infec Dis 17(4):359–361.

12. World Health Organization (2016) Report of the 16th meeting of the strategic and technical advisory group for tuberculosis Available at: https://www.who.int/publications/m/item/report-of-the-16th-meeting-of-the-strategic-and-technical-advisory-group-for-tb.

13. Abuali MM, Katariwala R, LaBombardi VJ (2012) A comparison of the Sensititre® MYCOTB panel and the agar proportion method for the susceptibility testing of Mycobacterium tuberculosis. Eur J Clin Micro Infect Dis 31(5):835–839.

14. Lee J, et al. (2014) Sensititre MYCOTB MIC plate for testing mycobacterium tuberculosis susceptibility to first-and second-line drugs. Antimicrob Agent Chemo 58(1):11–18.

15. Yu X, et al. (2016) Sensititre W MYCOTB MIC plate for drug susceptibility testing of Mycobacterium tuberculosis complex isolates. Int J Tuberc Lung Dis 20(3):329–334.

16. Xia H, et al. (2017) Assessment of a 96-well plate assay of quantitative drug susceptibility testing for mycobacterium tuberculosis complex in China. PLoS One 12(1):1–12.

17. Rancoita PM V., et al. (2018) Validating a 14-Drug Microtiter Plate Containing Bedaquiline and Delamanid for Large-Scale Research Susceptibility Testing of Mycobacterium tuberculosis. Antimicrob Agents Chemother 62(9):e00344–18.

18. Ruesen C, et al. (2018) Linking minimum inhibitory concentrations to whole genome sequence-predicted drug resistance in Mycobacterium tuberculosis strains from Romania. Sci Rep 8(1):1–8.

19. World Health Organization (2018) Rapid Communication: Key changes to treatment of multidrug-and rifampicin-resistant tuberculosis (MDR/RR-TB).

20. European Committee for Antimicrobial Susceptibility Testing (2017) MIC distributions and epidemiological cut-off value (ECOFF) setting. (EUCAST SOP 10.0):1–17.

21. The CRyPTIC Consortium (2022) https://github.com/fowler-lab/cryptic-ecoffs.

22. Ismail NA, et al. (2020) Epidemiological cut-offs for Sensititre susceptibility testing of Mycobacterium tuberculosis: interpretive criteria cross validated with whole genome sequencing. Sci Rep 10(1):1013.

23. Lipworth S, et al. (2019) SNP-IT tool for identifying subspecies and associated lineages of Mycobacterium tuberculosis complex. Emerg Infect Dis 25(3):482–488.

24. Gagneux S (2018) Ecology and evolution of Mycobacterium tuberculosis. Nat Rev Microbiol 16(4):202–213.

25. Turnidge J, Kahlmeter G, Kronvall G (2006) Statistical characterisation of bacterial wild-type MIC value distributions and the determination of epidemiological cut-off values. Clin Microbiol Infect 12(5):418–425.

26. Amemiya T (1973) Regression Analysis when the Dependent Variable Is Truncated Normal. Econometrica 41(6):997.

27. Hurd M (1979) Estimation in Truncated Samples when there is Heteroscedasticity. J Econom 11:247–258.

28. Ghodousi A, et al. (2019) Isoniazid Resistance in Mycobacterium tuberculosis Is a Heterogeneous Phenotype Composed of Overlapping MIC Distributions with Different Underlying Resistance Mechanisms. Antimicrob Agent Chemo 63(7):524157.

29. Ahmad S, Mokaddas E, Al-Mutairi N, Eldeen HS, Mohammadi S (2016) Discordance across phenotypic and molecular methods for drug susceptibility testing of drug-resistant Mycobacterium tuberculosis isolates in a low TB incidence country. PLoS One 11(4):1–16.

30. Farhat MR, et al. (2016) Gyrase Mutations Are Associated with Variable Levels of Fluoroquinolone Resistance in Mycobacterium tuberculosis. J Clin Microbiol 54(3):727–733.

31. Moore DAJ, et al. (2006) Microscopic-observation drug-susceptibility assay for the diagnosis of TB. New Eng J Med 355(15):1539–1550.

32. Clinical and Laboratory Standards Institute (2018) M23 - Development of in vitro susceptibility testing criteria and quality control parameters,(5th edition) (Wayne, PA, ISBN 1562388428).

33. Clinical and Laboratory Standards Institute (2018) M62 - Performance Standards for Susceptibility Testing of Mycobacteria, Nocardia spp., and Other Aerobic Actinomycetes (1st edition) (Wayne, PA, ISBN 9781684400270).

34. World Health Organization (2012) Updated interim critical concentrations for first-line and second-line DST.

35. World Health Organization (2018) Technical Report on critical concentrations for drug susceptibility testing of medicines used in the treatment of drug-resistant tuberculosis Available at: http://apps.who.int/iris/bitstream/handle/10665/260470/WHO-CDS-TB-2018.5-eng.pdf;jsessionid=07E7DB76974BC66918C6262AE55A733B?sequence=1.

36. World Health Organization (2021) Technical report on critical concentrations for drug susceptibility testing of isoniazid and the rifamycins (rifampicin, rifabutin and rifapentine).

37. Schön T, et al. (2009) Evaluation of wild-type MIC distributions as a tool for determination of clinical breakpoints for Mycobacterium tuberculosis. J Antimicrob Chemother 64(4):786–793.

38. Juréen P, et al. (2010) Wild-type MIC distributions for aminoglycoside and cyclic polypeptide antibiotics used for treatment of Mycobacterium tuberculosis infections. J Clin Microbiol 48(5):1853–1858.

39. Schön T, et al. (2011) Wild-type distributions of seven oral second-line drugs against Mycobacterium tuberculosis. Int J Tuberc Lung Dis 15(4):502–509.

40. Köser CU, Maurer FP, Kranzer K (2019) ‘Those who cannot remember the past are condemned to repeat it’: Drug-susceptibility testing for bedaquiline and delamanid. Int J Infect Dis:2017–2020.

41. Schena E, et al. (2016) Delamanid susceptibility testing of Mycobacterium tuberculosis using the resazurin microtitre assay and the BACTECTM MGITTM 960 system. J Antimicrob Chem 71(6):1532–1539.

42. Ismail NA, et al. (2018) Defining Bedaquiline Susceptibility, Resistance, Cross-Resistance and Associated Genetic Determinants: A Retrospective Cohort Study. EBioMedicine 28:136–142.

43. Kaniga K, et al. (2020) Validation of Bedaquiline Phenotypic Drug Susceptibility Testing Methods and Breakpoints: a Multilaboratory, Multicountry Study. J Clin Microbiol 58(4):1–10.

44. Ismail NA, et al. (2020) A Multimethod, Multicountry Evaluation of Breakpoints for Bedaquiline Resistance Determination. Antimicrob Agent Chemo 64(9):e00479–20.

45. Kahlmeter G, et al. (2020) Re: In the name of common sense: EUCAST breakpoints and potential pitfalls. National dissemination of EUCAST guidelines is a shared responsibility. Clin Microbiol Infect 26:1692–1693.

46. Schön T, et al. (2019) Standards for MIC testing that apply to the majority of bacterial pathogens should also be enforced for Mycobacterium tuberculosis complex. Clin Microbiol Infect 25(4):403–405.

47. Farhat MR, et al. (2019) Rifampicin and rifabutin resistance in 1003 Mycobacterium tuberculosis clinical isolates. J Antimicrob Chemother 74(6):1477–1483.

48. Shi W (2021) Activity of pyrazinamide against mycobacterium tuberculosis at neutral pH in PZA-S1 minimal medium. Antibiotics 10(8). doi:10.3390/antibiotics10080909.

49. Michael A, Kelman T, Pitesky M (2020) Overview of quantitative methodologies to understand antimicrobial resistance via minimum inhibitory concentration. Animals 10(8):1–17.

50. Kahlmeter G, Turnidge J, Brown D (2018) EUCAST General Consultation on “Considerations in the numerical estimation of epidemiological cutoff values”.

51. Zabeti H, Dexter N, Libbrecht M, Chindelevitch L (2020) An interpretable classification method for predicting drug resistance in M. tuberculosis. bioRxiv doi:101101/20200531115741. doi:10.1101/2020.05.31.115741.

52. Grazian C (2019) Estimating MIC distributions and cutoffs through mixture models_: an application to establish M. Tuberculosis resistance. bioRxiv doi:101101/643429. doi:10.1101/643429.

53. Hunt M, Letcher B, Hall MB, Lima L, Iqbal Z (2021) Minos: principled variant adjudication and joint genotyping using genome graphs (in preparation).

54. Hunt M (2021) Clockwork: Pipelines for processing bacterial sequence data (Illumina only) and variant calling. Available at: https://github.com/iqbal-lab-org/clockwork.

55. Walker TM, et al. (2014) Assessment of Mycobacterium tuberculosis transmission in Oxfordshire, UK, 2007-12, with whole pathogen genome sequences: An observational study. Lancet Resp Med 2(4):285–292.

56. Fowler PW (2021) Tuberculosis AMR catalogues in a standard grammar. Available at: https://github.com/oxfordmmm/tuberculosis_amr_catalogues.

57. Fowler PW (2020) gumpy: genetics with Numpy. Available at: https://github.com/oxfordmmm/gumpy.

58. Fowler PW (2021) piezo: predicting the effect of a genetic mutation on an antibiotic. Available at: https://github.com/oxfordmmm/piezo.

59. Dotson DL, Seyler SL, Linke M, Gowers RJ, Beckstein O (2016) datreant: persistent, Pythonic trees for heterogeneous data. Proc 15th Python Sci Conf, eds Benthall S, Rostrup S, pp 51–56.

60. Fowler PW, et al. (2018) Automated detection of bacterial growth on 96-well plates for high-throughput drug susceptibility testing of Mycobacterium tuberculosis. Microbiology 164(12):1522–1530.

61. Fowler PW (2020) AMyGDA. Available at: https://github.com/philipwfowler/amygda.

62. Fowler PW, et al. (2021) BashTheBug□ a crowd of volunteers reproducibly and accurately measure the minimum inhibitory concentrations of 13 antitubercular drugs from photographs of 96-well broth microdilution plates. bioRxiv doi:101101/20210720453060.

63. McKinney W (2010) Data Structures for Statistical Computing in Python. Proceedings of the 9th Python in Science Conference, ed Millman S van der W and J, pp 51–56.

64. Harris CR, et al. (2020) Array programming with NumPy. Nature 585(7825):357–362.

65. Hunter JD (2007) Matplotlib: A 2D Graphics Environment. Comput Sci Eng 9(3):90–95.

