## Supplemental Material for "Epidemiological cutoff values for a 96-well broth microdilution plate for high-throughput research antibiotic susceptibility testing of *M. tuberculosis*"

*see list at end of this document

| Laboratory | Plate Design Location | Total | UKMYC5 | UKMYC6 |
|---|---|---|---|---|
| African Health Research Institute | South Africa | 124 | 0 | 124 |
| Brazil | Brazil | 368 | 68 | 300 |
| Centers for Disease Control and Prevention | United States | 136 | 1 | 135 |
| Centre for Tuberculosis, NICD | South Africa | 1912 | 895 | 1017 |
| Chinese Center for Disease Control and Prevention | China | 2768 | 1409 | 1359 |
| Hinduja Hospital and Foundation for Medical Research Mumbai | India | 4992 | 2327 | 2665 |
| Institute of Microbiology and Laboratory Medicine | Germany | 1773 | 104 | 1669 |
| Oxford University Clinical Research Unit | Vietnam | 1509 | 679 | 830 |
| Public Health Sweden | Sweden | 457 | 100 | 357 |
| San Raffaele Scientific Institute | Italy | 2367 | 1134 | 1233 |
| TORCH | South Africa | 147 | 0 | 147 |
| Taiwan Centers for Disease Control | Taiwan | 171 | 171 | 0 |
| Universidad Peruana Cayetano Heredia | Peru | 3452 | 1077 | 2375 |
| University of Capetown | South Africa | 461 | 0 | 461 |
| Total | | 20637 | 7965 | 12672 |

Table S1: Total number of samples collected by laboratory, split by microtitre plate design. This table can be reproduced online[1].

| Country | Samples |
|---|---|
| India | 4992 |
| Peru | 3451 |
| China | 2767 |
| South Africa | 2638 |
| Italy | 1769 |
| Vietnam | 1509 |
| Germany | 1133 |
| Sweden | 457 |
| Pakistan | 408 |
| Brazil | 368 |
| Nepal | 302 |
| Turkmenistan | 256 |
| Taiwan | 171 |
| Belarus | 165 |
| United States | 136 |
| Ukraine | 30 |
| Kyrgyzstan | 28 |
| Algeria | 25 |
| Tajikistan | 19 |
| Kazakhstan | 4 |
| Somalia | 3 |
| Unknown | 1 |
| South Georgia and the South Sandwich Islands | 1 |
| New Caledonia | 1 |
| Chile | 1 |
| Slovenia | 1 |
| Japan | 1 |
| Total | 20,637 |

Table S2: Total number of samples grouped by country where collected[1]. Some laboratories only collected samples from within the country where they are based whilst others collected samples from several countries, hence this list is longer than Table S1.

| Species | Number of isolates |
|---|---|
| M. tuberculosis | 12348 |
| M. bovis BCG | 7 |
| M. orygis | 4 |
| M. bovis bovis | 2 |
| M. bovis caprae | 1 |

Table S3: All isolates contained a species belonging to the Mycobacterium complex[1], as determined by SNP-IT[2]. Since this was done by inspecting the genetics, this was only possible for isolates which had their whole genomes sequenced.

| Lineage | Number of isolates | Proportion (%) |
|---|---|---|
| Lineage 1 | 692 | 5.6 |
| Lineage 2 | 4358 | 35.3 |
| Lineage 3 | 1065 | 8.6 |
| Lineage 4 | 6227 | 50.4 |
| Lineage 6 | 6 | 0.0 |

Table S4: The majority of *M. tuberculosis* isolates belonged to either lineage 2 or 4[1], as determined by SNP-IT[2].

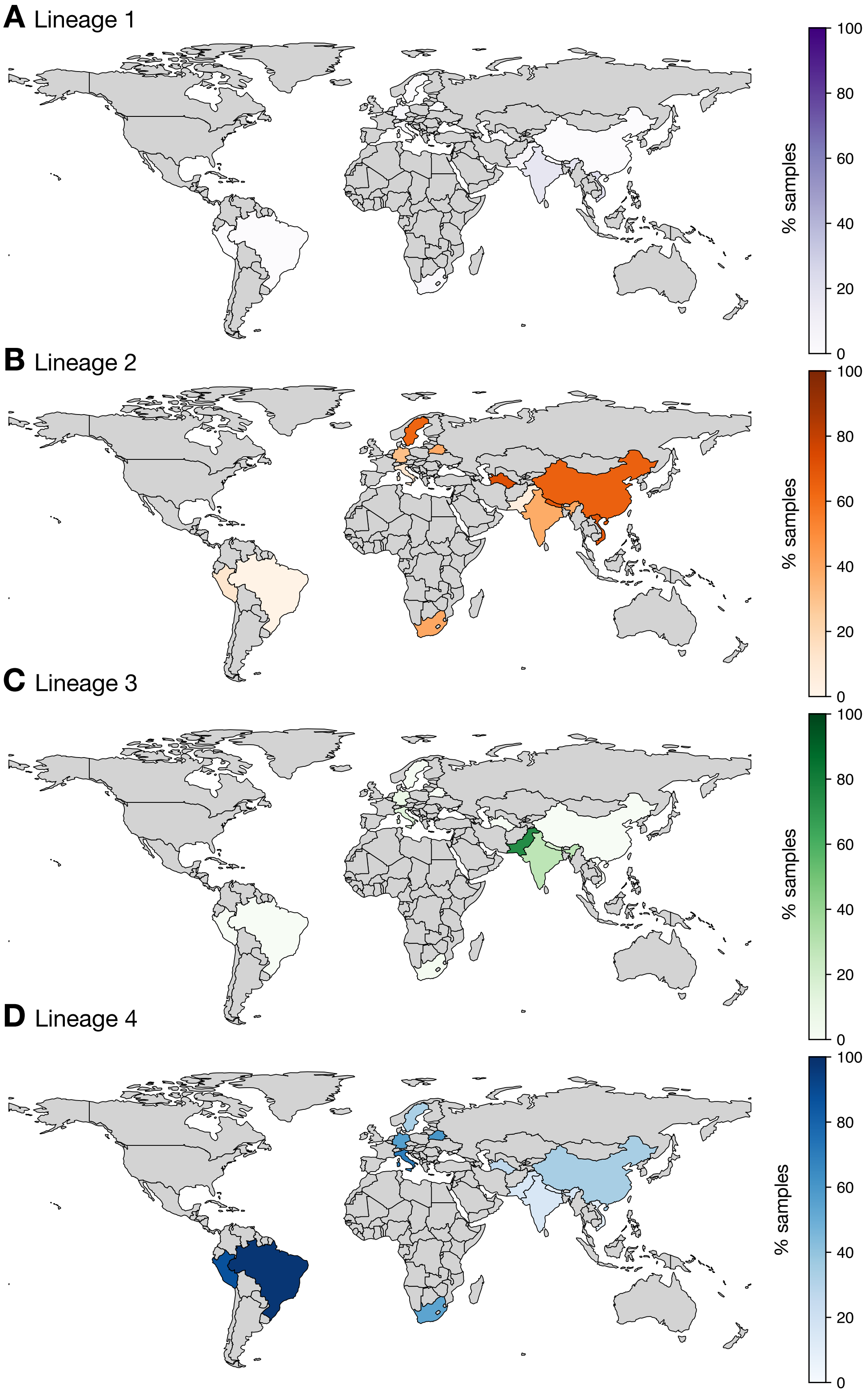

Figure S1: Choropleths showing the proportion of Lineages 1-4 in the isolates containing *M. tuberculosis* collected from each country[1]. Only countries where more than 100 samples were collected are shown and Lineage 6 is excluded due to the small number of samples.

| Lineage<br>Country | Lineage 1 | Lineage 2 | Lineage 3 | Lineage 4 | Total |
|---|---|---|---|---|---|
| Algeria | 0 | 0 | 0 | 25 | 25 |
| Belarus | 0 | 40 | 0 | 62 | 102 |
| Brazil | 1 | 8 | 0 | 334 | 343 |
| China | 0 | 702 | 1 | 371 | 1074 |
| Germany | 25 | 218 | 60 | 400 | 703 |
| India | 261 | 581 | 425 | 239 | 1506 |
| Italy | 52 | 148 | 179 | 909 | 1288 |
| Japan | 0 | 0 | 0 | 1 | 1 |
| Kyrgyzstan | 0 | 25 | 0 | 3 | 28 |
| Nepal | 5 | 137 | 34 | 21 | 197 |
| Pakistan | 18 | 21 | 300 | 64 | 403 |
| Peru | 1 | 339 | 0 | 2303 | 2643 |
| Slovenia | 0 | 0 | 0 | 1 | 1 |
| Somalia | 0 | 1 | 0 | 1 | 2 |
| South Africa | 61 | 858 | 55 | 1171 | 2145 |
| South Georgia and the South Sandwich Islands | 0 | 0 | 0 | 1 | 1 |
| Sweden | 6 | 271 | 6 | 141 | 424 |
| Tajikistan | 0 | 15 | 0 | 4 | 19 |
| Turkmenistan | 0 | 85 | 1 | 32 | 118 |
| Ukraine | 0 | 21 | 0 | 9 | 30 |
| Vietnam | 262 | 888 | 4 | 134 | 1288 |
| Total | 692 | 4358 | 1065 | 6227 | 12342 |

Table S5: Distribution of *M. tuberculosis* lineages by country where collected[1]. For clarity only isolates with Lineages 1-4 are shown.

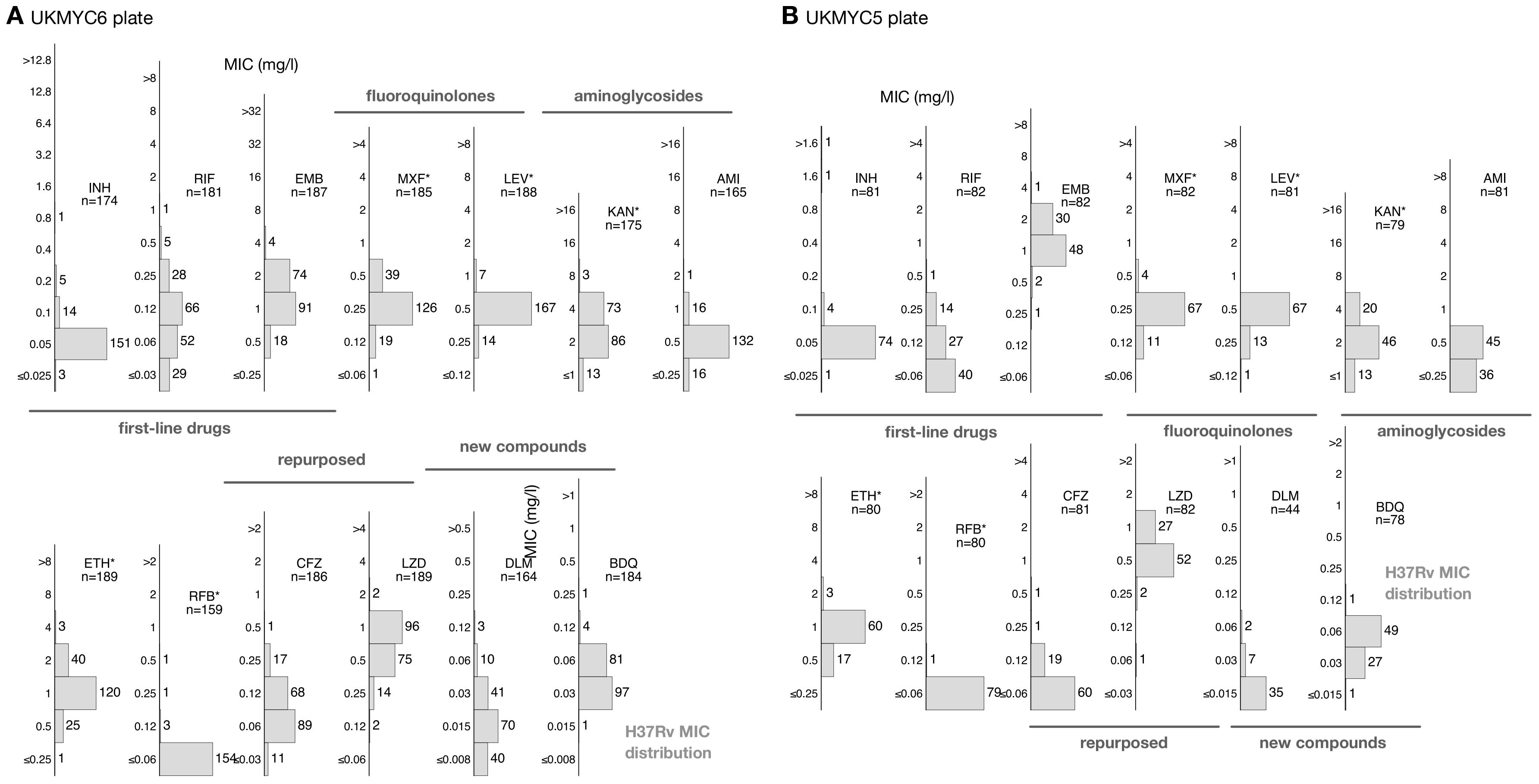

Figure S2: MIC distributions for the H37Rv reference strain on the (**A**) UKMYC6 and (**B**) UKMYC5 plates[1]. Data from eight and five laboratories, respectively, have been pooled. See Table S6 for the essential agreement by drug.

| Drug | Plate design | H37Rv mode MIC | Essential agreement | Number of isolates |
|---|---|---|---|---|
| INH | UKMYC6 | 0.05 mg/L | 94.9-96.6 % | (165-168)/174 |
| RIF | UKMYC6 | 0.12 mg/L | 80.7 % | 146/181 |
| EMB | UKMYC6 | 1 mg/L | 97.9 % | 183/187 |
| MXF | UKMYC6 | 0.25 mg/L | 99.5 % | 184/185 |
| LEV | UKMYC6 | 0.5 mg/L | 100 % | 188/188 |
| KAN | UKMYC6 | 2 mg/L | 90.9-98.3 % | (159-172/175 |
| AMI | UKMYC6 | 0.5 mg/L | 89.7-99.4 % | (148-164)/165 |
| ETH | UKMYC6 | 1 mg/L | 97.9 % | 185/189 |
| RFB | UKMYC6 | $\leq$ 0.06 mg/L | – | – |
| CFZ | UKMYC6 | 0.06 mg/L | 84.4-90.3 % | (157-168)/186 |
| LZD | UKMYC6 | 1 mg/L | 91.5 % | 173/189 |
| DLM | UKMYC6 | 0.015 mg/L | 67.7-92.1% | (111-151)/164 |
| BDQ | UKMYC6 | 0.03 mg/L | 97.3 % | 179/184 |
| INH | UKMYC5 | 0.05 mg/L | 96.3-97.5 % | (78-79)/81 |
| RIF | UKMYC5 | $\leq$ 0.06 mg/L | – | – |
| EMB | UKMYC5 | 1 mg/L | 97.6 % | 80/82 |
| MXF | UKMYC5 | 0.25 mg/L | 100 % | 82/82 |
| LEV | UKMYC5 | 0.5 mg/L | 98.8 % | 81/82 |
| KAN | UKMYC5 | 2 mg/L | 83.6-100 % | (66-79)/79 |
| AMI | UKMYC5 | 0.5 mg/L | 55.6-100 % | (45-81)/81 |
| ETH | UKMYC5 | 1 mg/L | 100% | 80/80 |
| RFB | UKMYC5 | $\leq$ 0.06 mg/L | – | – |
| CFZ | UKMYC5 | $\leq$ 0.06 mg/L | – | – |
| LZD | UKMYC5 | 0.5 mg/L | 98.8 % | 81/82 |
| DLM | UKMYC5 | $\leq$ 0.015 mg/L | – | – |
| BDQ | UKMYC5 | 0.06 mg/L | 98.7 % | 77/78 |

Table S6: The proportion of strains that lie within 1 dilution of the mode MIC of the standard H37Rv reference strain varies between drugs. Data for both plate designs are shown separately. For some of the drugs the mode is different on the two plates (LZD, BDQ) – the MIC distributions for these drugs are narrow with the majority of isolates contained within two doubling dilutions and hence it is difficult to define the mode. For other drugs the mode lies one well above the lowest dilution (INH, KAN, AMI, CFZ, DLM) on one or both of the plate designs, thereby introducing ambiguity into the essential agreement (EA). In these cases, the EA is calculated as a range that either includes or excludes the isolates in the bottom well. For yet other drugs, the mode lies in the lowest dilution (CFZ, RIF, RFB, DLM); the dilution range for CFZ, RIF and DLM was extended to lower values in the UKMYC6 plate design which partially resolved this issue. Five and eight laboratories contributed variable numbers of H37Rv readings to the UKMYC5 and UKMYC6 datasets, respectively. This study was not designed to determine the quality control ranges for H37Rv on the UKMYC series of plates, however this data could be used to guide future plate designs and experiments.

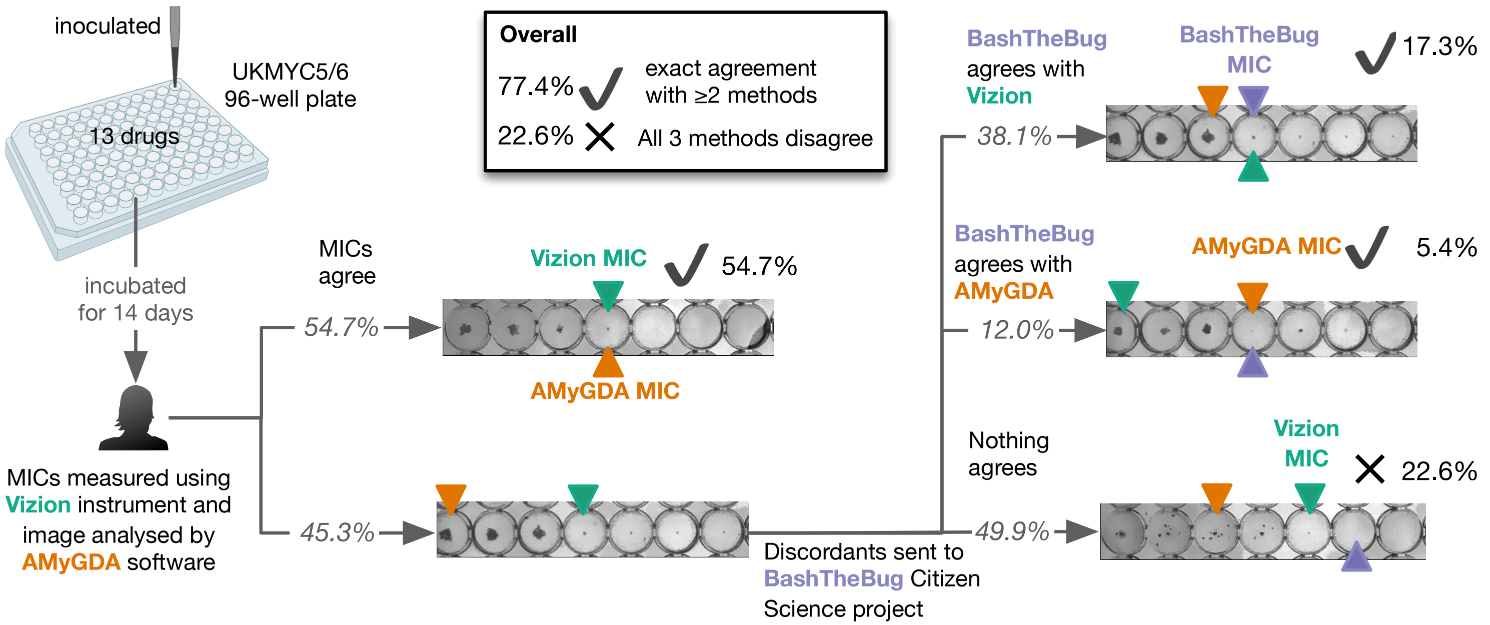

Figure S3: In our quality assurance workflow each minimum inhibitory concentration (MIC) was measured by two or three independent methods. (**A**) Following culture in a MGIT tube (Methods), a sample was inoculated onto a UKMYC plate. (**B**) After 14 days incubation, each plate was placed in a Thermo Fisher Sensititre Vizion Digital MIC Viewing System and the MICs were read by a trained laboratory scientist and stored in a central database[3]. A photograph of the plate was then taken using the Vizion instrument and the image uploaded to the same central database. (**C**) Each image was subsequently analysed centrally using the AMyGDA plate-reading software specifically developed for this purpose[4]. If the two MICs for a drug were identical (55% of all measurements), the process halts and that measurement is annotated as passing the control quality process. (**D**) If the MICs were different, an image of that drug doubling dilution series was uploaded to BashTheBug, a citizen science project. At least 11 different volunteers then measured the MIC and a consensus was taken. (**E**) Of the images sent to BashTheBug, the consensus MIC agreed with either the Vizion or AMyGDA reading in 39% and 12% of cases (Fig. S2). This resulted in a further 23% of the measurements passing the quality control process. (**F**) For the remaining 22% of measurements, all three MICs differed. These data were excluded from analyses.

| Antibiotic | Passed QA (%) | Failed QA (%) |
|---|---|---|
| AMI | 81.1 | 18.9 |
| BDQ | 78.6 | 21.4 |
| CFZ | 73.3 | 26.7 |
| DLM | 76.0 | 24.0 |
| EMB | 67.1 | 32.9 |
| ETH | 81.0 | 19.0 |
| INH | 88.7 | 11.3 |
| KAN | 84.4 | 15.6 |
| LEV | 71.5 | 28.5 |
| LZD | 68.3 | 31.7 |
| MXF | 63.5 | 36.5 |
| RFB | 91.7 | 8.3 |
| RIF | 80.6 | 19.4 |
| Average | 77.4 | 22.6 |

Table S7: The proportion of readings for each antibiotic where at least two methods agreed on the result and therefore have passed the quality control process. Cases where all three methods disagree fail the process. Only measurements where an image is available, a result was determined by the laboratory scientist and, if required, a consensus had been returned by the BashTheBug volunteers are included.

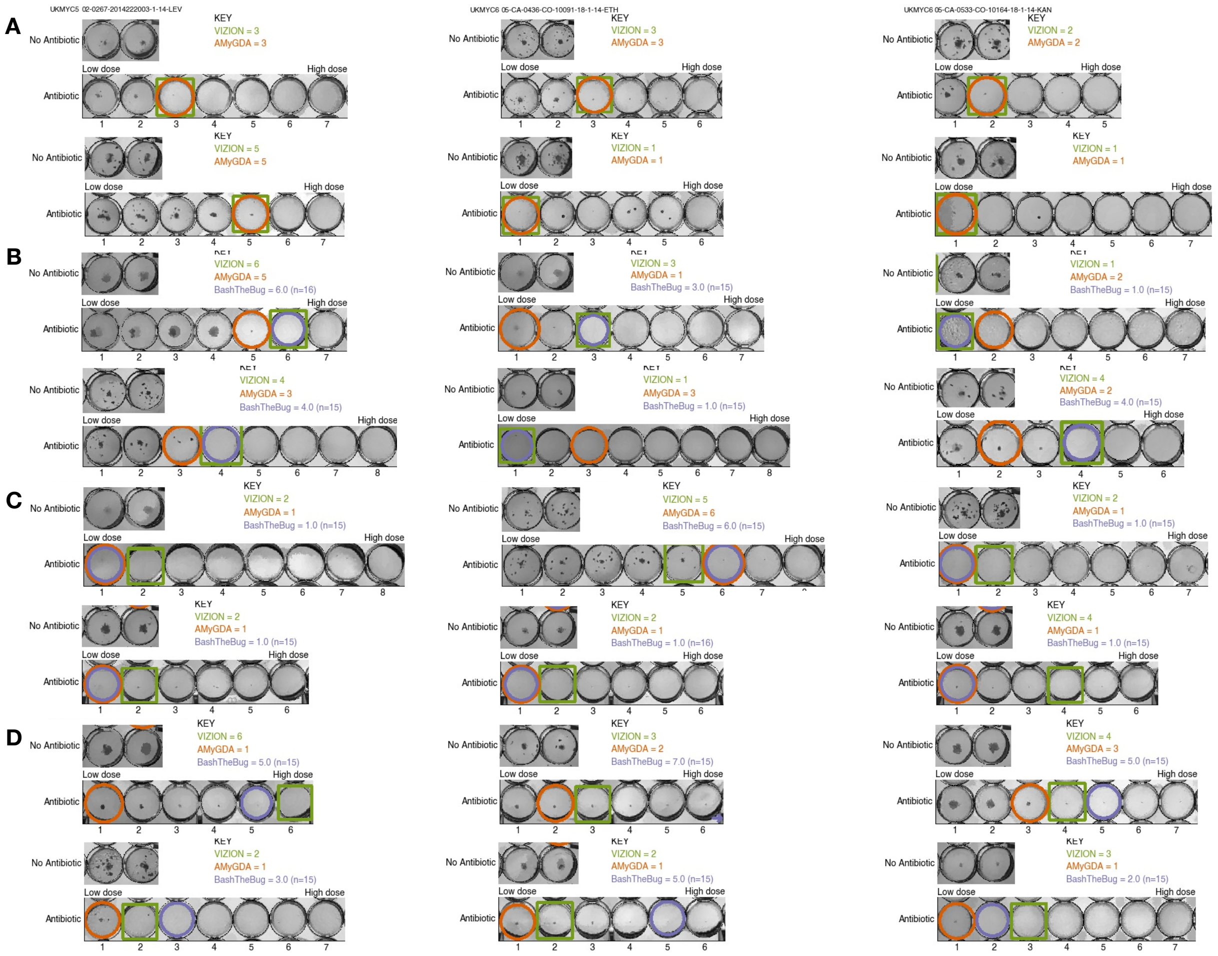

Figure S4: (Related to Figure S3) Randomly selected example images from different parts of the quality control workflow. (**A**) Six examples where the MIC measured by the laboratory scientist using the Vizion instrument and the AMyGDA software agree. (**B**) Six examples where the MIC measured by the laboratory scientist using the Vizion instrument and the consensus measured by the BashTheBug citizen scientists agree. In four cases AMyGDA assessed a lower MIC than the other methods as it incorrectly did not detect small growth in wells. In the remaining two cases, AMYGDA called a higher MIC since it incorrectly assessed growth due to artefacts. (**C**) Six examples where the MIC measured by AMyGDA and the BashTheBug consensus agree. The Vizion measurement appears to be either simply incorrect or the operator has assessed very small dots as growth, which is contrary to the CRyPTIC standard operating procedure. (**D**) Six examples where all three methods yield different MICs. These are a mixture of cases where e.g. all three methods call consecutive MICs and the tendency of the citizen scientists to be conservative in their assessment of growth can be seen.

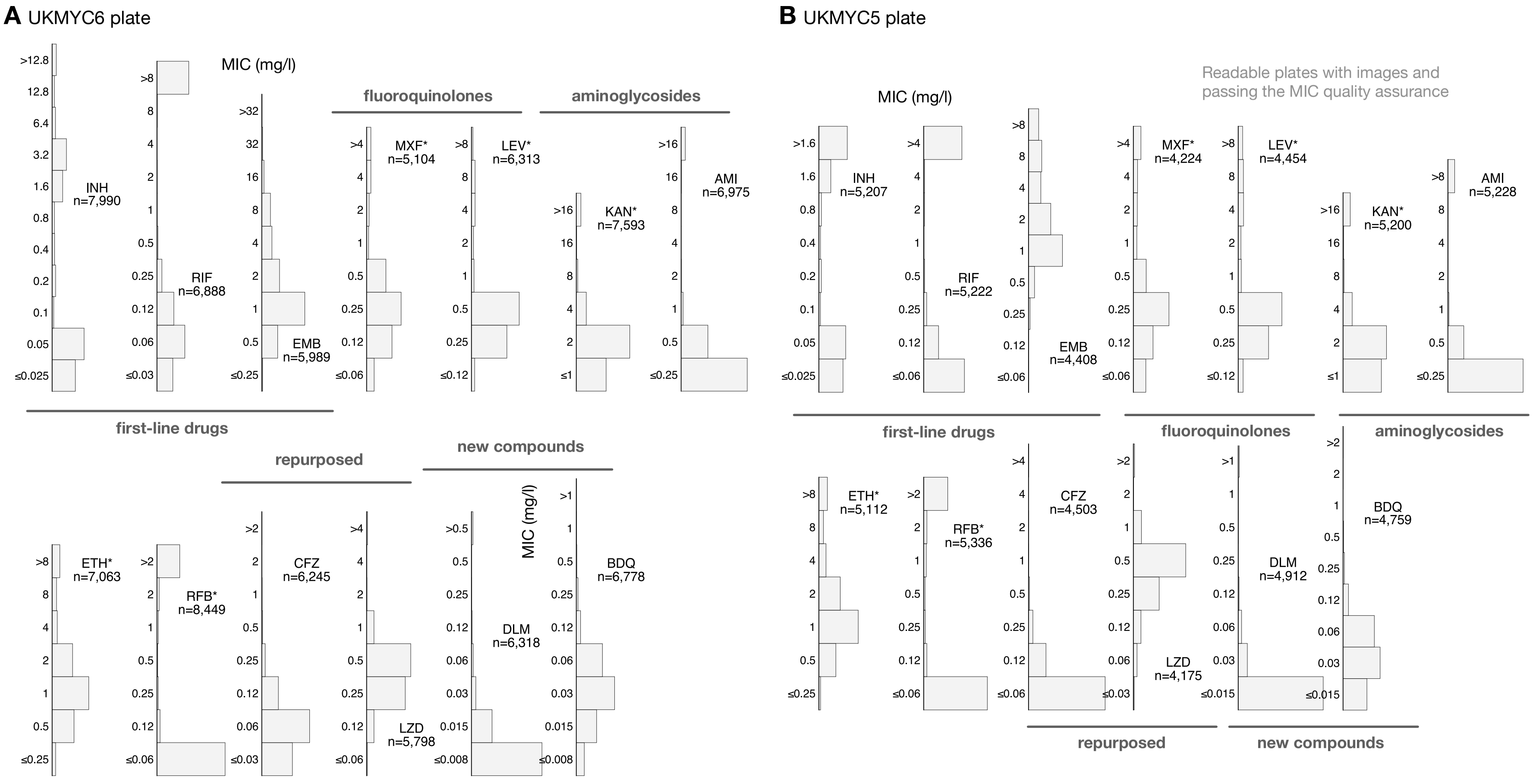

Figure S5: The histograms of the MICs, by drug, that passed the quality assurance process for the (**A**) UKMYC6 and (**B**) UKMYC5 plates[1]. Each MIC reading has therefore been confirmed by at least two independent methods. See the separate Supplemental CSV file for the numerical data.

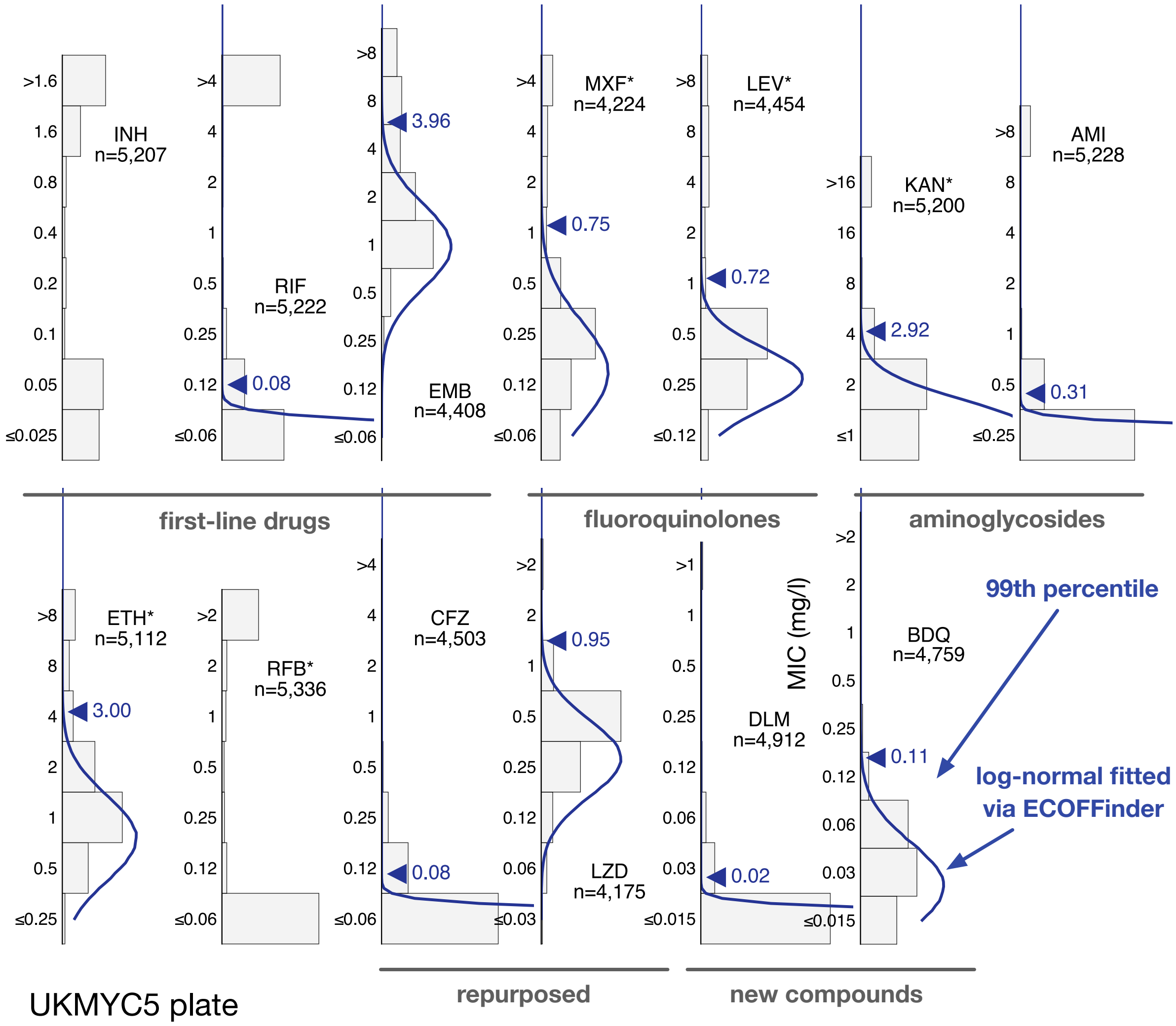

Figure S6: (Related to Figure 2) The MIC histograms for the 13 antibiotics on the UKMYC5 plate. Only MICs which have been confirmed by two independent measurement methods are shown. ECOFFinder was used to fit a log-normal distribution to each histogram; this is drawn in blue and the resulting 99th percentile is labelled. ECOFFinder was unable to fit a log-normal to isoniazid (INH) and rifabutin (RFB). See the separate Supplemental CSV file for the numerical data.

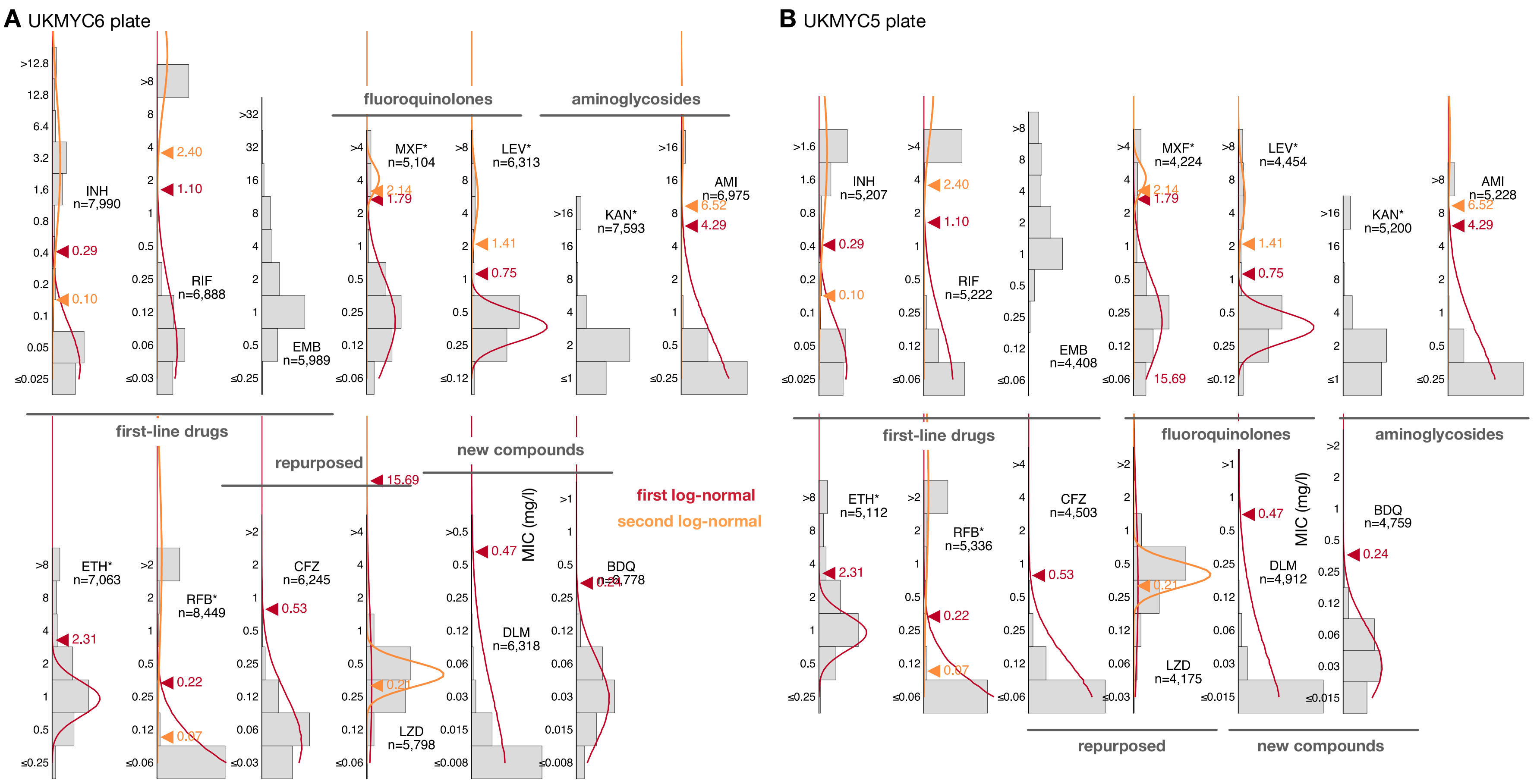

Figure S7: Two log-normal distributions were simultaneously fitted to the MIC histograms from each plate design for each drug using interval regression. The log-normal distribution with the smaller mean is coloured red and the other is coloured orange. The method fails to converge for kanamycin (KAN) and the variance of the second log-normal distribution is occasionally much larger than the range of the data (DLM, ETH)

| Laboratory | Plate Design Location | Total | UKMYC5 | UKMYC6 |
|---|---|---|---|---|
| Brazil | Brazil | 126 | 48 | 78 |
| Centre for Tuberculosis, NICD | South Africa | 592 | 333 | 259 |
| Chinese Center for Disease Control and Prevention | China | 463 | 463 | 0 |
| Hinduja Hospital and Foundation for Medical Research Mumbai | India | 365 | 365 | 0 |
| Institute of Microbiology and Laboratory Medicine | Germany | 439 | 8 | 431 |
| Oxford University Clinical Research Unit | Vietnam | 526 | 370 | 156 |
| San Raffaele Scientific Institute | Italy | 411 | 109 | 302 |
| Universidad Peruana Cayetano Heredia | Peru | 1039 | 354 | 685 |
| University of Capetown | South Africa | 342 | 0 | 342 |
| Total | | 4303 | 2050 | 2253 |

Table S8: Total number of genetically wild-type (gWT) isolates collected by laboratory, split by microtitre plate design.

| PLATEDESIGN<br>DRUG | UKMYC5 | UKMYC6 | Total |
|---|---|---|---|
| AMI | 1840 | 1667 | 3507 |
| BDQ | 1666 | 1631 | 3297 |
| CFZ | 1586 | 1414 | 3000 |
| DLM | 1687 | 1361 | 3048 |
| EMB | 1505 | 1354 | 2859 |
| ETH | 1832 | 1680 | 3512 |
| INH | 1813 | 1890 | 3703 |
| KAN | 1834 | 1790 | 3624 |
| LEV | 1587 | 1447 | 3034 |
| LZD | 1483 | 1297 | 2780 |
| MXF | 1477 | 1117 | 2594 |
| RFB | 1959 | 2119 | 4078 |
| RIF | 1810 | 1568 | 3378 |

Table S9: Number of genetically wild-type MICs by drug that have passed the quality assurance process. Note that only definite numerical MICs have been included.

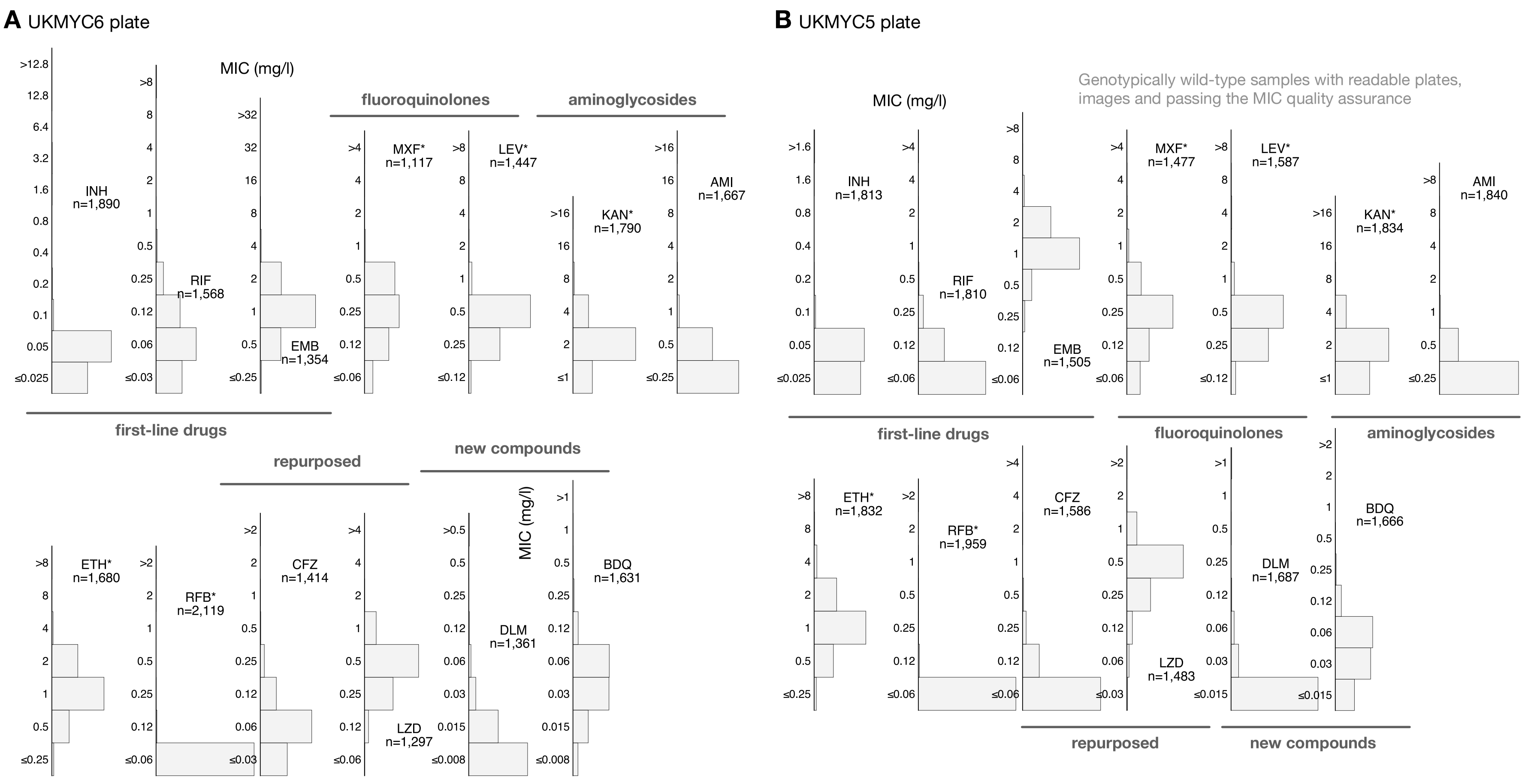

Figure S8: The histograms of all MICs that passed the quality assurance process and are *genotypically wild-type* for the (**A**) UKMYC6 and (**B**) UKMYC5 plates. Each MIC reading has therefore been confirmed by at least two independent methods.

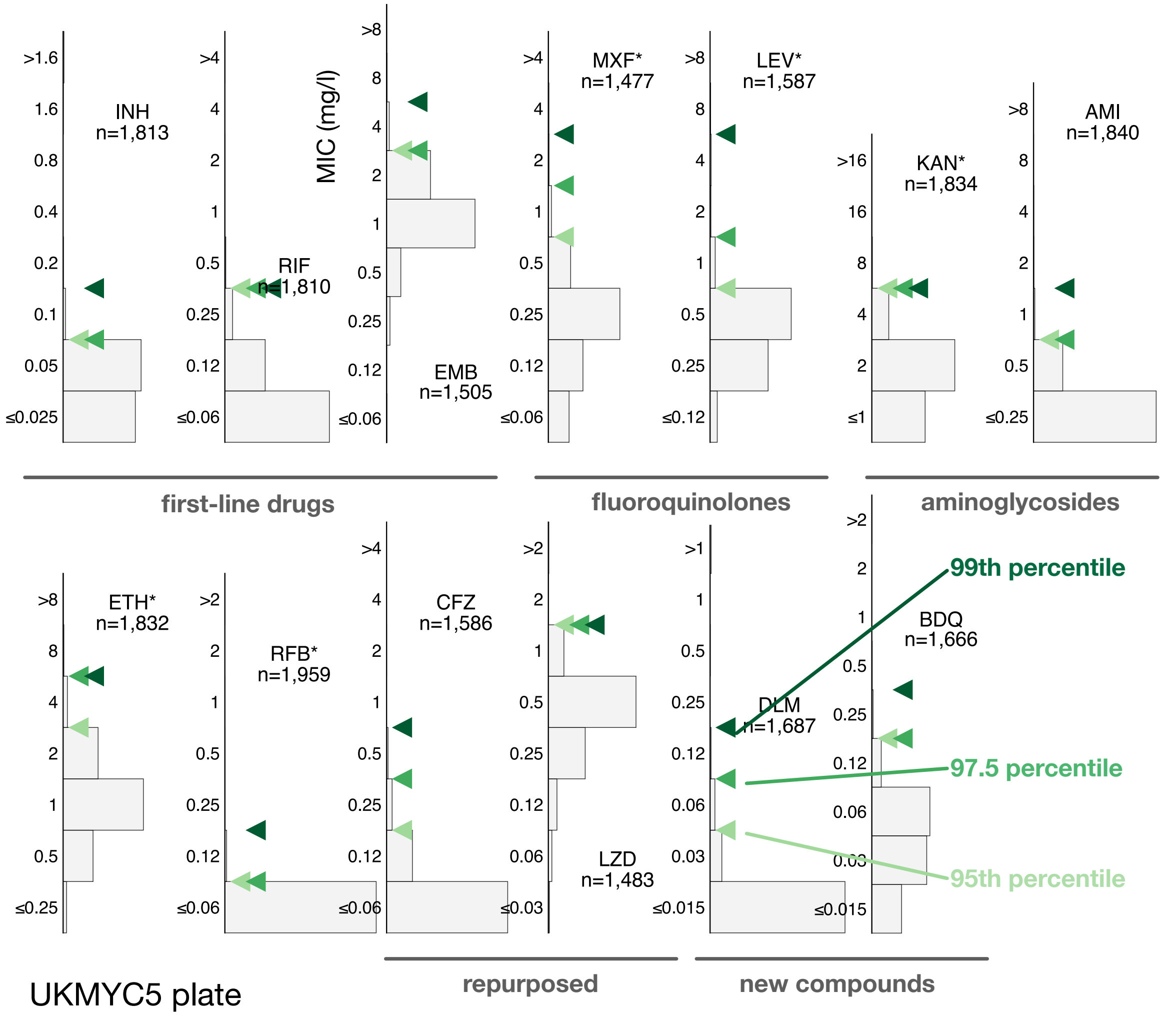

Figure S9: (Related to Figure 3) Direct measurement of ECOFF/ECVs from the gWT population on the UKMYC5 plate. To illustrate the sensitivity to the precise percentile used in the definition, the 95th, 97.5th and 99th percentiles are all labelled.

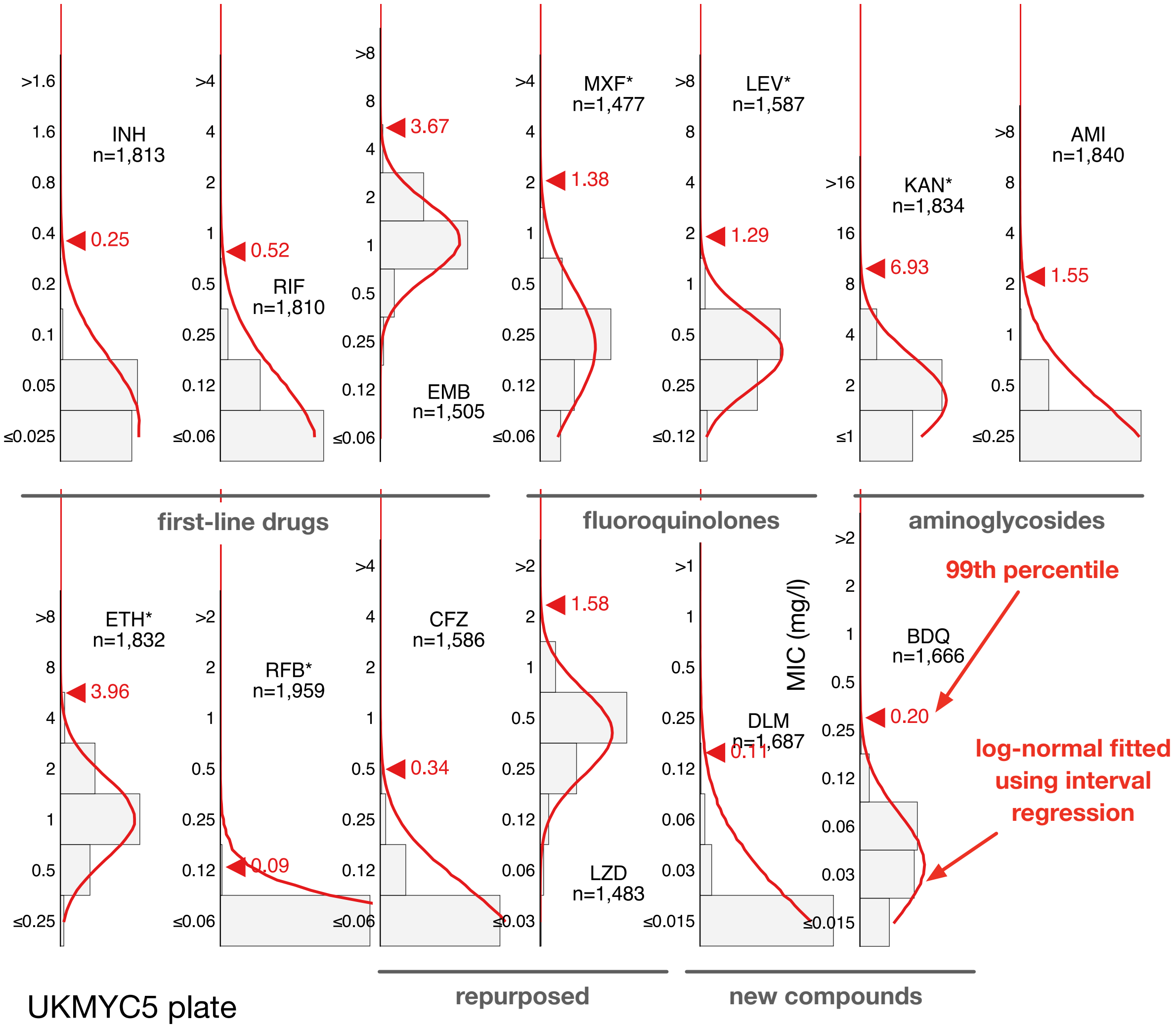

Figure S10: (Related to Figure 4) Interval regression is able to fit a log-normal distribution to the MIC histograms of the genetically wild-type isolates for the 13 drugs on the UKMYC5 plate. Data from both plate designs were considered simultaneously, hence the resulting distributions are those the algorithm considers to best describe both the UKMYC6 (Fig. 4) and UKMYC5 data sets.

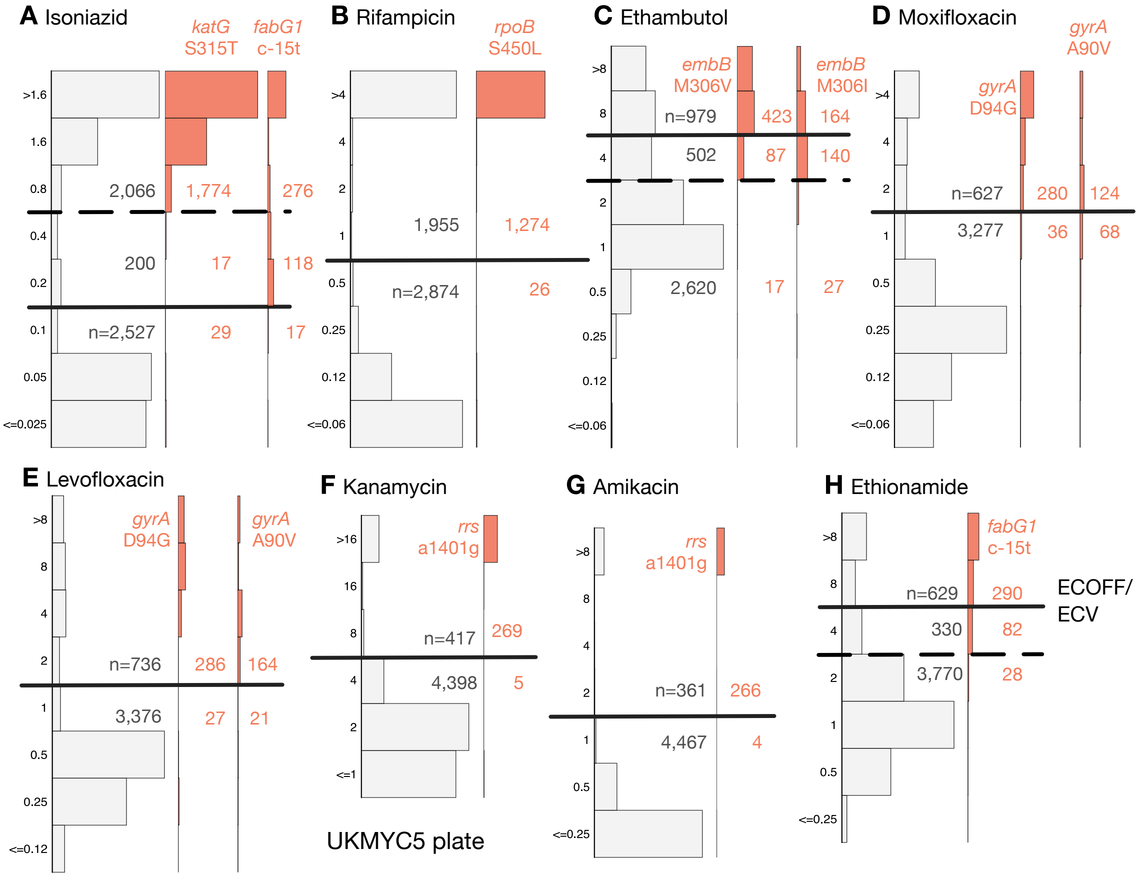

Figure S11: (Related to Figure 6) The MICs of isolates containing genetic variants known to confer resistance to different drugs tend to lie above the proposed ECOFF/ECV on the UKMYC6 plate. The number of isolates lying above and below the ECOFF/ECV is annotated. The dashed line indicates the margin of a proposed borderline category for isoniazid, ethambutol and ethionamide. The same analysis repeated on the UKMYC5 dataset is Figure 6 in the main body of the manuscript.

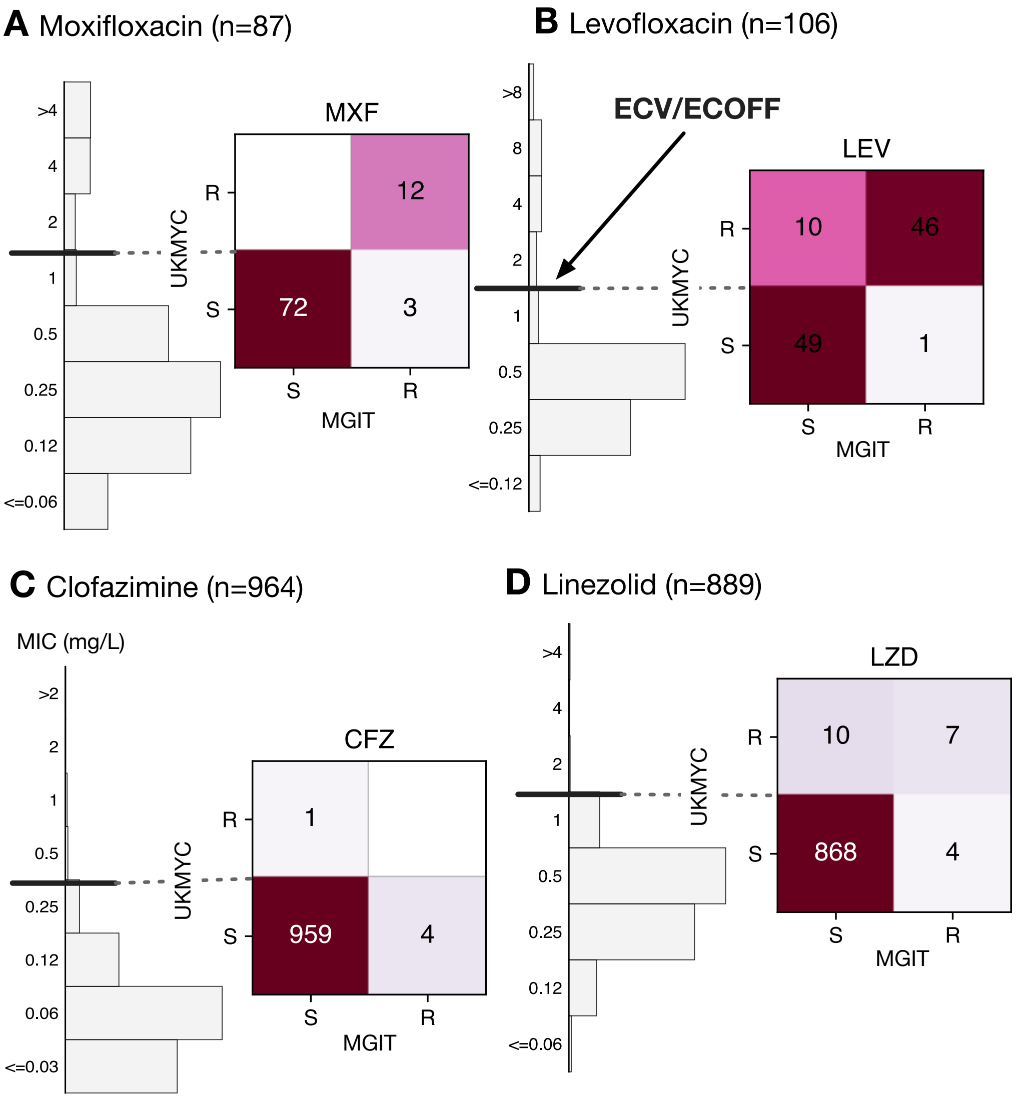

Figure S12: (Related to Figure 7) There is reasonable agreement between the phenotypes measured by the UK-MYC plates (assuming the ECOFF/ECVs) and the MGIT960. There are comparatively few isolates for the fluoroquinolones and very few resistant isolates for clofazimine and linezolid.

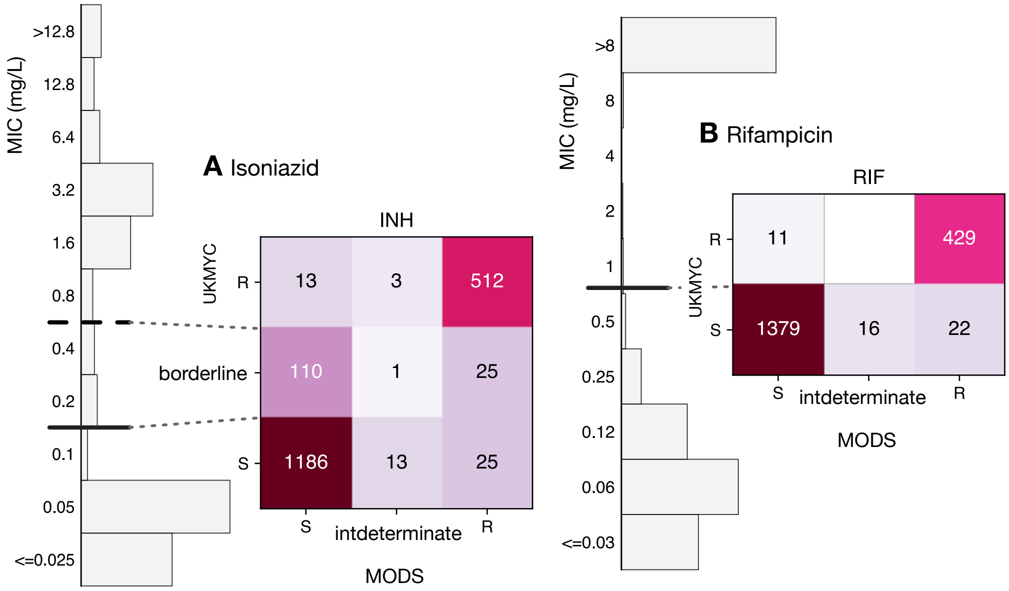

Figure S13: There is reasonable agreement between the phenotypes measured by the UKMYC plates (assuming the ECOFF/ECVs) and the MODS assay[5]. Only isoniazid and rifampicin were tested.

| Drug | Number of isolates | Sensitivity (%) | Specificity (%) | Categorical agreement (%) | MD (%) | VMD (%) |
|---|---|---|---|---|---|---|
| INH | 1516 | 93.4 | 97.0 | 95.0 | 6.6 | 3.0 |
| RIF | 1456 | 96.5 | 96.6 | 96.6 | 3.5 | 3.4 |
| EMB | 961 | 91.4 | 91.9 | 91.6 | 8.6 | 8.1 |
| MXF | 87 | 80.0 | NaN | NaN | 20.0 | NaN |
| LEV | 106 | 97.9 | 83.1 | 89.6 | 2.1 | 16.9 |
| KAN | 1262 | 76.2 | 99.1 | 96.4 | 23.8 | 0.9 |
| AMI | 1175 | 84.3 | 99.3 | 98.0 | 15.7 | 0.7 |
| ETH | 954 | 63.0 | 97.0 | 86.9 | 37.0 | 3.0 |
| LZD | 889 | 63.6 | 98.9 | 98.4 | 36.4 | 1.1 |
| CFZ | 964 | NaN | 99.9 | NaN | NaN | 0.1 |

Table S10: Comparing the binary phenotypes derived from a UKMYC plate results to MGIT960. The sensitivity and specificity were calculated ignoring any borderline categorisation.

| Drug | Number of isolates | Sensitivity (%) | Specificity (%) | Categorical agreement (%) | MD (%) | VMD (%) |
|---|---|---|---|---|---|---|
| INH | 1888 | 95.3 | 98.9 | 97.8 | 4.7 | 1.1 |
| RIF | 1857 | 95.1 | 99.2 | 98.2 | 4.9 | 0.8 |

Table S11: Comparing the binary phenotypes derived from a UKMYC plate results to MODS[5]. The sensitivity and specificity were calculated ignoring any borderline categorisation.

**Members of the CRyPTIC consortium**

Ivan Barilar[29], Simone Battaglia[1], Emanuele Borroni[1], Angela Pires Brandao[2,3], Alice Brankin[4], Andrea Maurizio Cabibbe[1], Joshua Carter[5], Daniela Maria Cirillo[1], Pauline Claxton[6], David A Clifton[4], Ted Cohen[7], Jorge Coronel[8], Derrick W Crook[4], Viola Dreyer[29], Sarah G Earle[4], Vincent Escuyer[9], Lucilaine Ferrazoli[3], Philip W Fowler[4], George Fu Gao[10], Jennifer Gardy[11], Saheer Gharbia[12], Kelen Teixeira Ghisi[3], Arash Ghodousi[1,13], Ana Luíza Gibertoni Cruz[4], Louis Grandjean[33], Clara Grazian[14], Ramona Groenheit[44], Jennifer L Guthrie[15,16], Wencong He[10], Harald Hoffmann[17,18], Sarah J Hoosdally[4], Martin Hunt[4,19], Zamin Iqbal[19], Nazir Ahmed Ismail[20], Lisa Jarrett[21], Lavania Joseph[20], Ruwen Jou[22], Priti Kambli[23], Rukhsar Khot[23], Jeff Knaggs[4,19], Anastasia Koch[24], Donna Kohlerschmidt[9], Samaneh Kouchaki[4,25], Alexander S Lachapelle[4], Ajit Lalvani[26], Simon Grandjean Lapierre[27], Ian F Laurenson[6], Brice Letcher[19], Wan-Hsuan Lin[22], Chunfa Liu[10], Dongxin Liu[10], Kerri M Malone[19], Ayan Mandal[28], Mikael Mansjö[44], Daniela Matias[21], Graeme Meintjes[24], Flávia de Freitas Mendes[3], Matthias Merker[29], Marina Mihalic[18], James Millard[30], Paolo Miotto[1], Nerges Mistry[28], David Moore[8,31], Kimberlee A Musser[9], Dumisani Ngcamu[20], Hoang Ngoc Nhung[32], Stefan Niemann[29,48], Kayzad Soli Nilgiriwala[28], Camus Nimmo[33], Nana Okozi[20], Rosangela Siqueira Oliveira[3], Shaheed Vally Omar[20], Nicholas Paton[34], Timothy EA Peto[4], Juliana Maira Watanabe Pinhata[3], Sara Plesnik[18], Zully M Puyen[35], Marie Sylvianne Rabodoarivelo[36], Niaina Rakotosamimanana[36], Paola MV Rancoita[13], Priti Rathod[21], Esther Robinson[21], Gillian Rodger[4], Camilla Rodrigues[23], Timothy C Rodwell[37,38], Aysha Roohi[4], David Santos-Lazaro[35], Sanchi Shah[28], Thomas Andreas Kohl[29], Grace Smith[12,21], Walter Solano[8], Andrea Spitaleri[1,13], Philip Supply[39], Utkarsha Surve[23], Sabira Tahseen[40], Nguyen Thuy Thuong Thuong[32], Guy Thwaites[4,32], Katharina Todt[18], Alberto Trovato[1], Christian Utpatel[29], Annelies Van Rie[41], Srinivasan Vijay[42], Timothy M Walker[4,32], A Sarah Walker[4], Robin Warren[43], Jim Werngren[44], Maria Wijkander[44], Robert J Wilkinson[26,45,46], Daniel J Wilson[4], Penelope Wintringer[19], Yu-Xin Xiao[22], Yang Yang[4], Zhao Yanlin[10], Shen-Yuan Yao[20], Baoli Zhu[47].

**Affiliations**

1. IRCCS San Raffaele Scientific Institute, Milan, Italy
2. Oswaldo Cruz Foundation, Rio de Janeiro, Brazil
3. Institute Adolfo Lutz, São Paulo, Brazil
4. University of Oxford, Oxford, UK
5. Stanford University School of Medicine, Stanford, USA
6. Scottish Mycobacteria Reference Laboratory, Edinburgh, UK
7. Yale School of Public Health, Yale, USA
8. Universidad Peruana Cayetano Heredia, Lima, Perú
9. Wadsworth Center, New York State Department of Health, Albany, USA
10. Chinese Center for Disease Control and Prevention, Beijing, China
11. Bill & Melinda Gates Foundation, Seattle, USA
12. UK Health Security Agency, London, UK

13. Vita-Salute San Raffaele University, Milan, Italy
14. University of New South Wales, Sydney, Australia
15. The University of British Columbia, Vancouver, Canada
16. Public Health Ontario, Toronto, Canada
17. SYNLAB Gauting, Munich, Germany
18. Institute of Microbiology and Laboratory Medicine, IMLred, WHO-SRL Gauting, Germany
19. EMBL-EBI, Hinxton, UK
20. National Institute for Communicable Diseases, Johannesburg, South Africa
21. Public Health England, Birmingham, UK
22. Taiwan Centers for Disease Control, Taipei, Taiwan
23. Hinduja Hospital, Mumbai, India
24. University of Cape Town, Cape Town, South Africa
25. University of Surrey, Guildford, UK
26. Imperial College, London, UK
27. Université de Montréal, Canada
28. The Foundation for Medical Research, Mumbai, India
29. Research Center Borstel, Borstel, Germany
30. Africa Health Research Institute, Durban, South Africa
31. London School of Hygiene and Tropical Medicine, London, UK
32. Oxford University Clinical Research Unit, Ho Chi Minh City, Viet Nam
33. University College London, London, UK
34. National University of Singapore, Singapore
35. Instituto Nacional de Salud, Lima, Perú
36. Institut Pasteur de Madagascar, Antananarivo, Madagascar
37. FIND, Geneva, Switzerland
38. University of California, San Diego, USA
39. Institut Pasteur de Lille, Lille, France
40. National TB Reference Laboratory, National TB Control Program, Islamabad, Pakistan

41. University of Antwerp, Antwerp, Belgium

42. University of Edinburgh, Edinburgh, UK

43. Stellenbosch University, Cape Town, South Africa

44. Public Health Agency of Sweden, Solna, Sweden

45. Wellcome Centre for Infectious Diseases Research in Africa, Cape Town, South Africa

46. Francis Crick Institute, London, UK

47. Institute of Microbiology, Chinese Academy of Sciences, Beijing, China

48. German Center for Infection Research (DZIF), Hamburg-Lübeck-Borstel-Riems, Germany

### References

1. The CRyPTIC Consortium (2022). https://github.com/fowler-lab/cryptic-ecoffs.

2. Lipworth S, Jajou R, De Neeling A, Bradley P, Van Der Hoek W, Maphalala G, Bonnet M, Sanchez-Padilla E, Diel R, Niemann S, Iqbal Z, Smith G, Peto T, Crook D, Walker T, Van Soolingen D (2019) *Emerging Infectious Diseases* 25:482–488.

3. Rancoita PMV, Cugnata F, Gibertoni Cruz AL, Borroni E, Hoosdally SJ, Walker TM, Grazian C, Davies TJ, Peto TEA, Crook DW, Fowler PW, Cirillo DM, Crook DW, Peto TEA, Walker AS, Hoosdally SJ, Gibertoni Cruz AL, Grazian C, Walker TM, Fowler PW, Wilson D, Clifton D, Iqbal Z, Hunt M, Smith EG, Rathod P, Jarrett L, Matias D, Cirillo DM, Borroni E, Battaglia S, Chiacchiaretta M, De Filippo M, Cabibbe A, Tahseen S, Mistry N, Nilgiriwala K, Chitalia V, Ganesan N, Papewar A, Rodrigues C, Kambli P, Surve U, Khot R, Niemann S, Kohl T, Merker M, Hoffmann H, Lehmann S, Plesnik S, Ismail N, Omar SV, Joseph L, Marubini E, Thwaites G, Thuy Thuong TN, Ngoc NH, Srinivasan V, Moore D, Coronel J, Solano W, He G, Zhu B, Zhou Y, Ma A, Yu P, Schito M, Claxton P, Laurenson I (2018) *Antimicrobial Agents and Chemotherapy* 62:e00344–18.

4. Fowler PW, Gibertoni Cruz AL, Hoosdally SJ, Jarrett L, Borroni E, Chiacchiaretta M, Rathod P, Lehmann S, Molodtsov N, Grazian C, Walker TM, Robinson E, Hoffmann H, Peto TEA, Cirillo DM, Smith GE, Crook DW (2018) *Microbiology* 164:1522–1530.

5. Moore DAJ, Evans CAW, Gilman RH, Caviedes L, Coronel J, Vivar A, Sanchez E, Piñedo Y, Saravia JC, Salazar C, Oberhelman R, Hollm-Delgado MG, LaChira D, Escombe AR, Friedland JS (2006) *New Eng J Med* 355:1539–1550.
